# The COVIDome Explorer Researcher Portal

**DOI:** 10.1101/2021.03.04.21252945

**Authors:** Kelly D. Sullivan, Matthew D. Galbraith, Kohl T. Kinning, Kyle Bartsch, Nik Levinsky, Paula Araya, Keith P. Smith, Ross E. Granrath, Jessica R. Shaw, Ryan Baxter, Kimberly R. Jordan, Seth Russell, Monika Dzieciatkowska, Julie A. Reisz, Fabia Gamboni, Francesca Cendali, Tusharkanti Ghosh, Andrew A. Monte, Tellen D. Bennett, Michael G. Miller, Elena W.Y. Hsieh, Angelo D’Alessandro, Kirk C. Hansen, Joaquin M. Espinosa

**Affiliations:** Linda Crnic Institute for Down Syndrome, University of Colorado Anschutz Medical Campus, Aurora, CO, USA; Department of Pediatrics, Section of Developmental Biology, University of Colorado Anschutz Medical Campus, Aurora, CO, USA; Department of Pharmacology, University of Colorado Anschutz Medical Campus, Aurora, CO, USA; Information Services, University of Colorado Anschutz Medical Campus, Aurora, CO, USA; Department of Immunology and Microbiology, University of Colorado Anschutz Medical Campus, Aurora, CO, USA; Data Science to Patient Value, University of Colorado Anschutz Medical Campus, Aurora, CO, USA; Department of Biochemistry and Molecular Genetics, University of Colorado Anschutz Medical Campus, Aurora, CO, USA; Department of Biostatistics and Informatics, Colorado School of Public Health, Aurora, CO, USA; Department of Emergency Medicine, University of Colorado Anschutz Medical Campus, Aurora, CO, USA; Department of Pediatrics, Sections of Informatics and Data Science and Critical Care Medicine, University of Colorado Anschutz Medical Campus, Aurora, CO, USA; Department of Pediatrics, Division of Allergy/Immunology, University of Colorado Anschutz Medical Campus, Aurora, CO, USA

**Keywords:** COVID-19, SARS, multi-omics, data portal, CRP, inflammation, infection

## Abstract

COVID-19 pathology involves dysregulation of diverse molecular, cellular, and physiological processes. In order to expedite integrated and collaborative COVID-19 research, we completed multi-omics analysis of hospitalized COVID-19 patients including matched analysis of the whole blood transcriptome, plasma proteomics with two complementary platforms, cytokine profiling, plasma and red blood cell metabolomics, deep immune cell phenotyping by mass cytometry, and clinical data annotation. We refer to this multidimensional dataset as the COVIDome. We then created the COVIDome Explorer, an online researcher portal where the data can be analyzed and visualized in real time. We illustrate here the use of the COVIDome dataset through a multi-omics analysis of biosignatures associated with C-reactive protein (CRP), an established marker of poor prognosis in COVID-19, revealing associations between CRP levels and damage-associated molecular patterns, depletion of protective serpins, and mitochondrial metabolism dysregulation. We expect that the COVIDome Explorer will rapidly accelerate data sharing, hypothesis testing, and discoveries worldwide.

## INTRODUCTION

Throughout the course of the COVID-19 pandemic, researchers around the world have made significant progress in the understanding of diverse aspects of the condition, including the epidemiology of SARS-CoV-2 infection and the underlying molecular, cellular, and physiological processes dysregulated in COVID-19 patients. This included completion of sophisticated genetic, molecular, and cellular analyses, as well as launching of myriad clinical trials. In many instances the rapid pace of discoveries has been facilitated by the assembly of large collaborations. Another factor accelerating the pace of research is the widespread use of pre-print collections where papers under peer-review can be accessed freely ahead of publication. However, we posit that the speed of research is being hampered by the lack of widely accessible, analysis-ready public datasets that could be analyzed in real time by experts and non-experts alike. Although great progress has been made in publication policy in terms of ensuring that the data fueling published discoveries are made accessible through public data repositories, most datasets remain inaccessible to broad audiences and can be downloaded and re-analyzed only by experts. In order to further accelerate research at a global scale, we created a multidimensional dataset derived from hospitalized COVID-19 patients versus COVID-19-negative controls, known as the COVIDome dataset, and made it readily accessible through a user-friendly platform, the COVIDome Explorer researcher portal.

The COVIDome dataset includes demographics and clinical data, along with matched analysis of the whole blood transcriptome via RNAseq (measuring 16,000+ RNAs), analysis of the plasma proteome by complementary SOMAscan® assays (measuring 5000+ epitopes), mass-spectrometry (400+ abundant proteins), and multiplexed cytokine profiling (80+ immune modulatory factors), analysis of the plasma and red blood cell metabolomes by mass-spectrometry, deep immune phenotyping by mass cytometry (measuring 50+ immune cell types), and seroconversion assays. All datasets are publicly accessible through a user-friendly, analysis-ready researcher portal dubbed the COVIDome Explorer (www.covidome.org). In this manuscript, we describe how the datasets were generated and analyzed, and explain how to use the COVIDome Explorer for rapid hypotheses testing, hypothesis generation, and real-time discoveries by experts and non-experts. We illustrate the prowess of the COVIDome dataset by completing a multi-omics analysis of biosignatures associated with varying levels of C-reactive protein (CRP), a clinical marker of poor prognosis in COVID-19 (Liu et al., 2020; Xu et al., 2020). This analysis revealed that high CRP levels associate with damage-associated molecular patterns (DAMPs), depletion of key members of the serpin family of serine protease inhibitors, and metabolic changes indicative of mitochondrial dysfunction.

## RESULTS

### The COVIDome: a multi-omics dataset for the study of COVID-19

In order to investigate variations in the endotype of COVID-19 patients, we completed a multi-omics assessment of 105 research participants, including 73 hospitalized COVID-19 patients versus 32 COVID-19-negative controls (**Figure 1A**). The demographics and clinical characteristics of this cohort are described in **Supp. File 1**. All COVID-19-positive participants were hospitalized due to moderate symptoms, but none had developed severe clinical disease requiring ICU admission at the time of blood collection. COVID-19 positivity was defined from results of PCR and/or antibody testing within 14 days of the research blood draw. Blood samples were analyzed by a matched multi-omics assessment of the transcriptome via RNAseq of whole blood, plasma proteomics using two alternative platforms [mass spectrometry (MS) and SOMAscan®], cytokine profiling using multiplexed immunoassays for 80+ immune factors using Meso Scale Discovery (MSD) assays, plasma and red blood cell (RBC) metabolomics via mass spectrometry, immune cell phenotyping via mass cytometry (MC), and seroconversion assays for detection of antibodies against SARS-CoV-2 nucleocapsid and spike polypeptides (**Figure 1A**). Importantly, all datasets were generated from different fractions of the exact same blood draw from each research participant, which enables effective cross-platform analyses.

**Figure 1.**
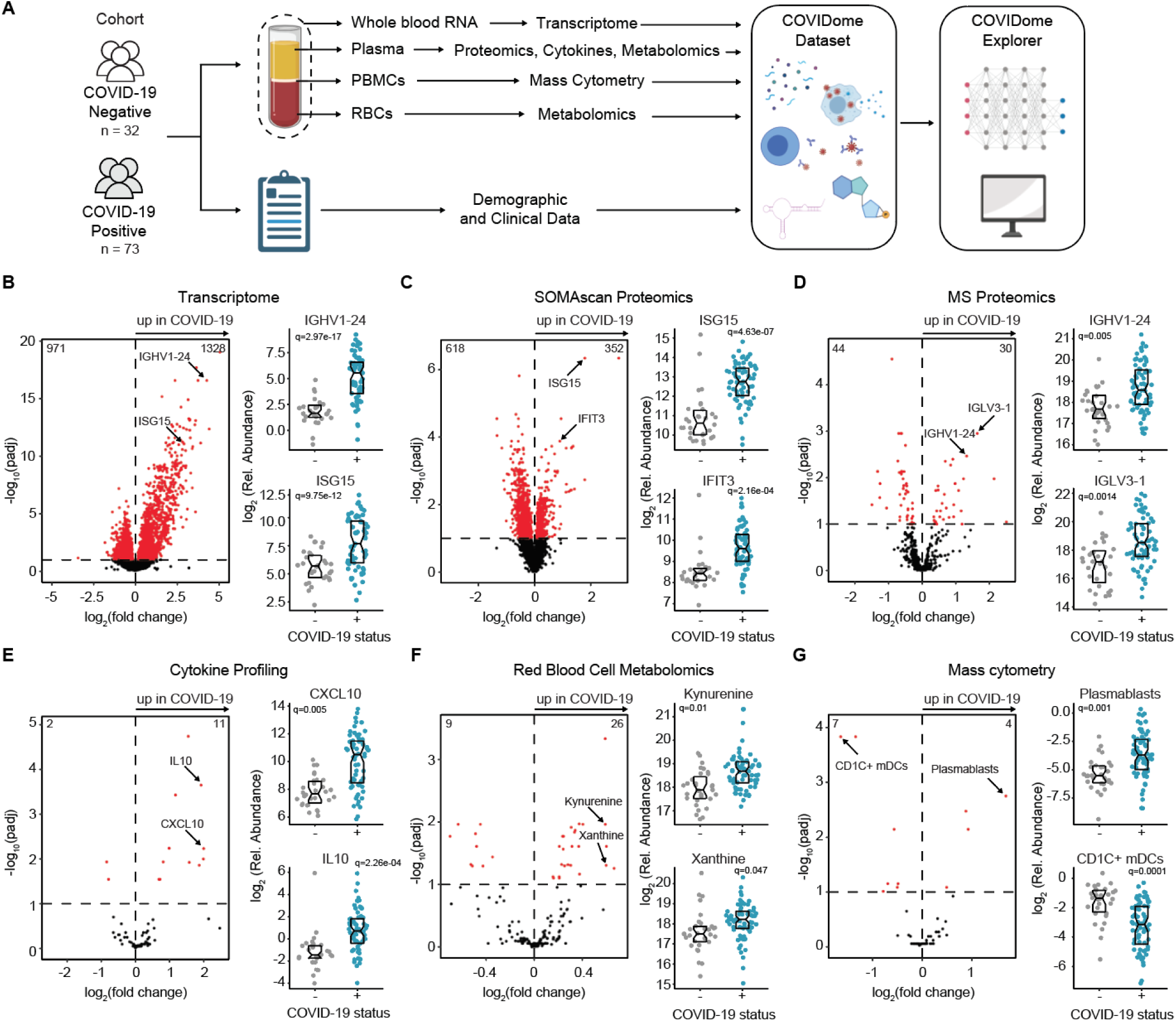
The COVIDome Dataset. **A.** Schematic of experimental approach. Blood samples were collected and processed for multi-omic analysis. Created with graphic elements from BioRender.com. **B-G. Left**, volcano plot indicating the impact of COVID-19, and **Right**, sina plots with boxes indicating median and interquartile range of representative features for (**B**) whole blood transcriptome, (**C**) plasma SOMAscan® proteomics, (**D**) plasma mass spectrometry (MS) proteomics, (**E**) plasma cytokine profiling, (**F**) red blood cell mass spectrometry metabolomics, and (**G**) mass cytometry of peripheral blood mononuclear cells (PBMCs). In the volcano plots, the vertical dashed midlines indicate no change in COVID-19 patients versus controls and the horizontal dashed lines indicate the statistical cut off of q<0.1 (FDR10). The numbers at the top left and right of each volcano indicate the number of features passing the statistical cut-off. In the sina plots, q values were calculated with DeSeq2 (transcriptome, adjusted for age and sex) or mixed linear models adjusting for age and sex (all other datasets).

To generate the transcriptome dataset, whole blood was collected in PAXgene RNA tubes, RNA extracted and subjected to next generation sequencing (see Methods). Analysis of the transcriptome dataset using DeSeq2 (Love et al., 2014) identified 2299 differentially expressed genes (DEGs) in the bloodstream of the COVID-19 patients (**Figure 1B**). Examples of significantly upregulated DEGs include specific immunoglobulin sequences (e.g. *IGHV1-24*), indicative of seroconversion, as well as interferon-stimulated genes (e.g. *Interferon-stimulated gene 15*, *ISG15*), indicative of an antiviral transcriptional response. An interactive volcano plot similar to that in **Figure 1B** enabling real time data visualization can be found in the Transcriptome dashboard of the COVIDome Explorer at https://covidome.shinyapps.io/Transcriptome/. DeSeq2 results can be found in **Supp. File 2**.

To generate the SOMAscan® proteomics dataset, plasma was analyzed with SOMAmer® technology to measure the abundance of 5000+ epitopes corresponding to 3000+ unique proteins (see Methods). Using a linear model adjusting for age and sex, we identified 970 differentially abundant epitopes in the plasma of COVID-19 patients (**Figure 1C**). Examples of significantly upregulated proteins include many ISGs, such as ISG15 and IFIT3 (Interferon Induced Protein with Tetratricopeptide Repeats 3) (**Figure 1C**). To generate the MS proteomics dataset, the same plasma aliquot used for SOMAscan® proteomics was analyzed by MS (see Methods). This approach enabled the quantification of 412 abundant proteins in plasma (**Figure 1D**). The MS proteomics dataset is highly complementary to the SOMAscan® proteomics dataset, as it enables detection of many abundant proteins for which SOMAmer® reagents are not available. For example, analysis of the MS proteomics dataset using a linear model adjusting for age and sex identified 74 differentially abundant proteins, including clear upregulation of immunoglobulin sequences not detected by the SOMAscan® but which were also detected as upregulated in the transcriptome dataset (e.g. IGHV1-24, IGLV3-1) (**Figure 1D**). Interactive volcano plots and box and whisker plots for the two proteomics datasets can be generated in the Proteome dashboard of the COVIDome Explorer at: https://covidome.shinyapps.io/Proteome/. Results of the mix linear models described here can be found in **Supp. File 3** (SOMAscan® proteomics) and **Supp. File 4** (MS proteomics).

To generate the cytokine profile dataset, the same aliquot of plasma used for the proteomics analyses was employed to measure the levels of a select list of immune modulatory factors via multiplexed immunoassays using MSD assays. A linear model adjusting for age and sex revealed many cytokines differentially abundant in the bloodstream of COVID-19 patients, such as CXCL10 (C-X-C motif chemokine ligand 10, interferon-inducible protein 10, IP10) and IL10 (Interleukin 10) (**Figure 1E**). Interactive volcano plots and box and whisker plots for this dataset can be generated in the Cytokine dashboard of the COVIDome Explorer at: https://covidome.shinyapps.io/Cytokines/. Results of the linear model for MSD data can be found in **Supp. File 5**.

To investigate metabolic dysregulation in COVID-19, we completed parallel targeted analyses of the RBC and plasma metabolomes using ultra-high pressure liquid chromatography coupled to mass spectrometry (UHPLC-MS) (see Methods). RBC and plasma metabolomic signatures inform about different metabolic and physiological processes, with both common and unique metabolites measured in each matrix. Analysis of the RBC metabolome revealed 35 differentially abundant metabolites in the COVID-19 patients, such as upregulation of kynurenine, a sign of activation of the IFN-inducible kynurenine pathway of tryptophan catabolism (Thomas et al., 2020), and xanthine, a sign of dysregulated purine metabolism (**Figure 1F**). Identical analysis of the plasma metabolome revealed many differentially abundant metabolites in COVID-19 patients, including kynurenine and xanthine as well (**Figure S1**). Interactive volcano plots and box and whisker plots for the two metabolomics datasets can be generated in the Metabolome dashboard of the COVIDome Explorer at: https://covidome.shinyapps.io/Metabolome/. Results of the linear models for metabolomics can be found in **Supp. File 6** (RBC metabolomics) and **Supp. File 7** (Plasma metabolomics).

Lastly, we completed a comprehensive map of peripheral immune cell lineages using MC, which enabled the identification and curation of 100+ immune cell subsets (see Methods). Toward this end, we utilized peripheral blood mononuclear cells (PBMCs) purified by Ficoll gradient from the same blood draw used for all other datasets and stained them with a panel of 40 metal-couple antibodies designed to quantify many major and minor lymphoid and myeloid subsets (see Methods). In order to quantify differences in immune cell subsets within their parent lineage, we created 7 different immune maps, stemming from: 1) all live cells, 2) CD3+ T cells (all T cells), 3) CD4+ T cells, 4) CD8+ T cells, 5) CD19+ B cells, 6) CD11c+ monocytes (CD3-CD19-CD56-), and 7) CD1c+ myeloid dendritic cells (mDCs) (CD3-CD19-CD56-) (**Supp. File 8**). Using a linear model adjusting for age and sex, we identified many immune cell types with significantly different frequency among all live cells in COVID-19-positive patients, such as increased frequencies of plasmablasts and decreased frequencies of CD1c+ mDCs (**Figure 1G**). Interactive volcano plots and box and whisker plots for the 7 immune maps can be generated in the Immune Maps dashboard of the COVIDome Explorer at: https://covidome.shinyapps.io/ImmuneMaps/. Results of the mix linear models for each immune lineage can be found in **Supp. File 8**.

In sum, the COVIDome dataset includes major data types for the study of diverse biological processes dysregulated in hospitalized COVID-19 patients.

### The COVIDome Explorer: an online portal for real-time data analysis, visualization, and sharing

In order to facilitate quick and broad access to the COVIDome dataset, we created a user-friendly online portal, dubbed the COVIDome Explorer, which can be accessed online at covidome.org (see overview in **Figure 2**).

**Figure 2.**
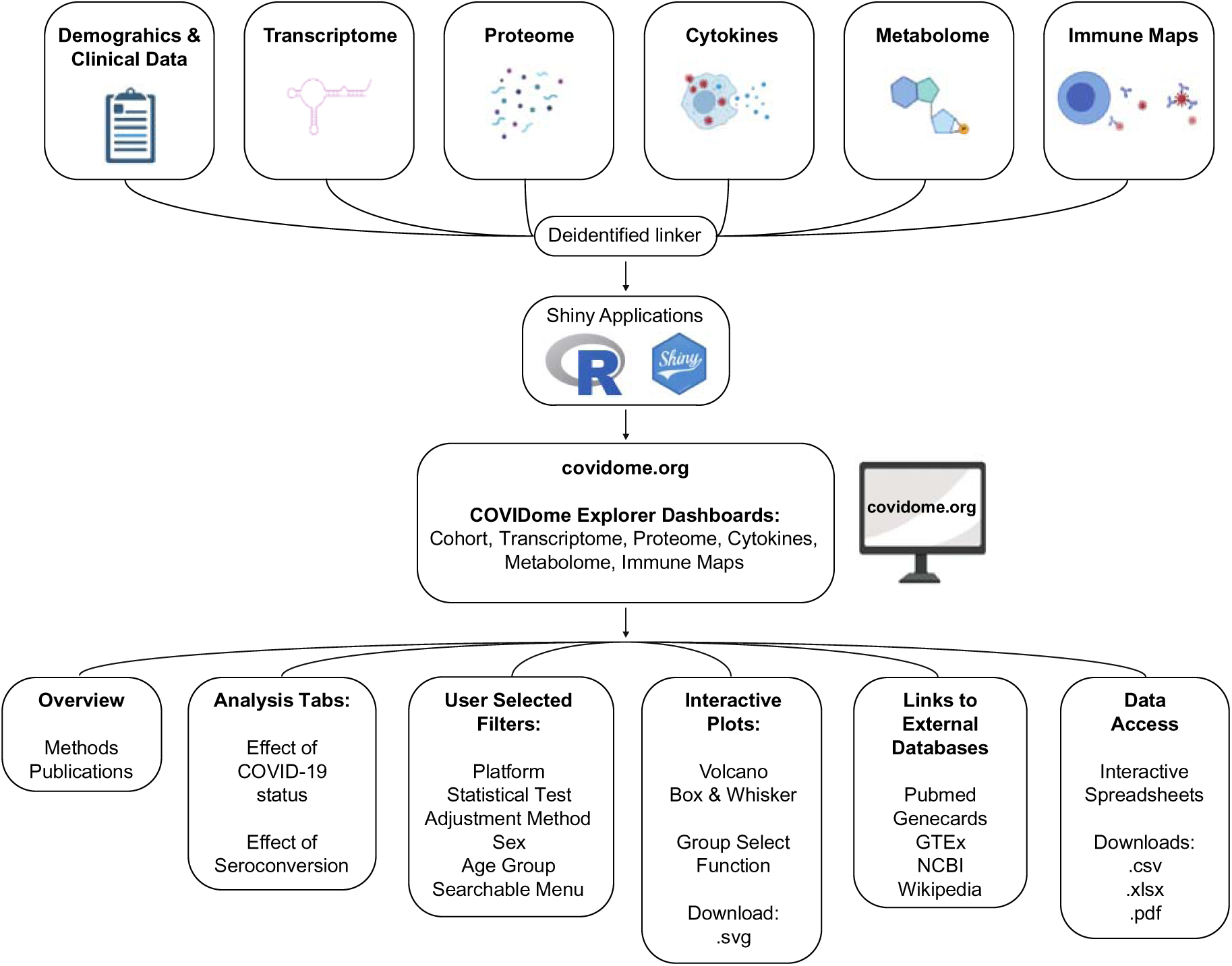
The COVIDome Researcher Portal. Schematic illustrating the design of the COVIDome Explorer researcher portal and its various functionalities. Created with graphic elements from BioRender.com.

After data curation and quality control, each of the COVIDome datasets was linked at the sample level with a unique identifier, enabling cross-referencing among platforms. Then, each of the datasets were imported into applications developed using R, R Studio, and the R-based web application framework Shiny. Each application includes custom-developed features that enable rapid query, visualization, and download of data, in an interactive environment (see Methods). The COVIDome Explorer hosts six dashboards: Cohort, Transcriptome, Proteome, Cytokines, Metabolome, and Immune Maps. Each dashboard runs within its own isolated and protected environment, hosted on the cloud-based Platform-as-a-Service (PaaS) environment “shinyapps.io.” When a user navigates to a specific dashboard via URL, individual instances of the Shiny application are instantly deployed to the shinyapps.io hosting platform, allowing for interaction and analysis throughout the duration of the user’s session. The Cohort dashboard is a simple description of the research cohort involved. The other dashboards are organized in a similar fashion and present similar options for analysis. This similarity allows users to become familiar with one dashboard, and then rapidly adapt to the use of the other dashboards. Each the five analytical dashboards contains three tabs: Overview, Effect of COVID-19 status, and Effect of Seroconversion. The Overview tab provides a summary of the approach, a brief explanation on how to use the dashboards, and in some instances, links to data files that would guide users, such as catalogs of proteins, metabolites, cytokines and immune cells present in each dataset. The Overview tab also points to publications that provide further detail about the methodology employed. The ‘Effect of COVID-19 status’ tab enables users to investigate differences between the COVID-19-negative control cohort and COVID-19-positive patients. The ‘Effect of Seroconversion’ tab enables users to investigate differences among COVID-19 patients with low versus high titers of anti-SARS-CoV-2 antibodies. A detailed description of the metrics of seroconversion employed and the definition of ‘sero-low’ versus ‘sero-high’ groups can be found in (Galbraith et al., 2020).

Upon entry into a given dashboard, users must select from a menu of options before data can be displayed. For the Proteome and Metabolome dashboards the first option is the choice of Platform: Mass spectrometry versus SOMAscan® for the Proteome, Plasma versus Red Blood Cells for the Metabolome. For the Immune Maps dashboard, the first choice is to select one of 7 parent lineages: Live Cells, CD3+ T, CD4+ T, CD8+ T, CD19+ B cells, Monocytes, Myeloid DCs. Next, users can choose a statistical test (linear model with age and sex adjustment; Kolmogorov-Smirnov test, Student’s T test, or Wilcoxon Test) and an Adjustment Method for multiple hypotheses correction [None, Bonferroni, Benjamini-Hochberg (false discovery rate, FDR)]. Users with a pre-formed hypothesis in mind interested in searching for a specific feature of interest (e.g. specific mRNA, protein or immune cell type) may opt out of a multiple hypothesis adjustment method. In contrast, users exploring the data in an unbiased fashion should select an adjustment method to account for multiple hypotheses testing. Two other filters are sex (both, Male, Female) and Age (All, 21 & Over), which enable users to visualize all or a fraction of the dataset. At this stage, users can *‘Apply filters and generate plot’*, which would then lead to the appearance of an interactive Volcano plot displaying the results. Users can then ‘mouse over and click’ individual features in the Volcano plot to display a box and whisker plot for that specific feature. Alternatively, users can use the searchable menus to find a feature of interest. Once an individual feature has been selected, live links to external databases become available, including Pubmed, GeneCards, GTEx, NCBI and Wikipedia, thus allowing users to navigate away from the COVIDome Explorer and learn more about a gene, protein, cytokine, metabolite, or immune cell type of interest. Of note, both Volcano Plots and Box and Whisker Plots can be downloaded as scalable vector graphics (.svg) files.

In each dashboard, the data being visualized can be accessed through the ‘*Aggregated Data’* or ‘*Sample Level Data’* tabs, two distinct interactive spreadsheets. In these tabs, users can filter by fold change and p value, sort by any of the columns visible (e.g. gene/protein name, fold change, p value), and search for individual features. Users can then download the data as comma separated values file (.csv), Microsoft Excel spreadsheet (.xlsx), or pdf files.

Altogether, the COVIDome Explorer dashboards enable data access and analysis by a broad range of users with different degrees of bioinformatics and biostatistics literacy, from those simply interested in a group comparison for a single protein, to those interested in sophisticated off-line analyses of the downloaded datasets.

### CRP levels associate with damage-associated molecular patterns

To illustrate the utility of the matched multidimensional COVIDome datasets, we analyzed multi-omics biosignatures associated with varying levels of CRP, an acute phase protein whose elevation in circulation has been consistently associated with poor prognosis in COVID-19. Repeatedly, higher CRP levels at the time to hospitalization and/or rapid rise in CRP levels during hospitalization have been associated with increased probability of developing severe COVID-19 pathophysiology (Mousavi-Nasab et al., 2020; Mueller et al., 2020; Sharifpour et al., 2020). As expected, CRP is also elevated in our cohort of COVID-19 patients as measured by MS proteomics, as well as other acute phase proteins such as ferritin (FTL) (**Figure 3A**). In order to identify biosignatures associated with CRP levels among COVID-19-positive patients, we calculated Spearman correlations between CRP values measured by MS and all features in all COVIDome datasets, which revealed myriad mRNAs, proteins, and metabolites significantly associated with CRP levels (**Figure S2A-F**, **Supp. Files 9-15**). This analysis exercise confirmed known associations, such as positive correlations between CRP levels and the levels of serum amyloid proteins SAA1 and SAA2, the acute phase protein LBP (Lipopolysaccharide Binding Protein), and the cytokines IL6 and IL10 (**Figure 3B**, **Figure S2B**) (Jain et al., 2011). Notably, there were no significant associations between CRP levels and frequencies of immune cell types, neither among all live cells or within major lymphoid and myeloid lineages, with the sole exception of increase frequencies of inflammatory subsets of monocytes (**Figure S2G-H**, **Supp. File 15**).

**Figure 3.**
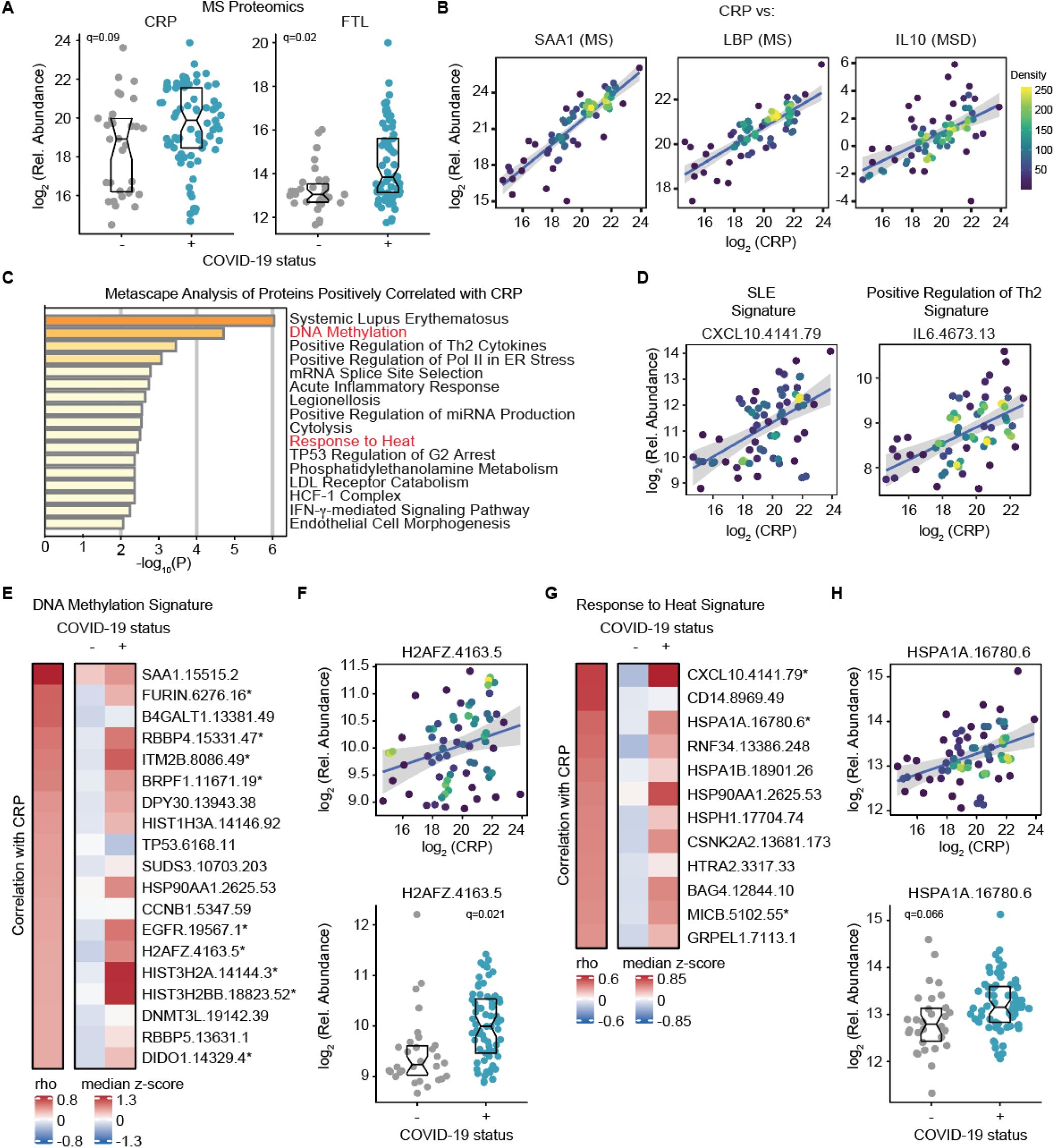
CRP levels correlate with damage associated molecular patterns. **A.** Sina plots showing values for immune factors correlated CRP levels comparing COVID-19-negative (-) to COVID-19-positive (+) patients. Data are presented as modified Sina plots with boxes indicating median and interquartile range. **B.** Scatter plots displaying correlations between CRP levels SAA1, LBP, and IL10. MS: mass spectrometry; MSD: Meso Scale Discovery assay. Points are colored by density; lines represent linear model fit with 95% confidence interval. **C.** Metascape pathway enrichment analysis of proteins detected by SOMAscan® proteomics that are significantly and positively correlated with CRP. **D.** Scatter plot displaying correlations between CRP levels and representative factors from the Systemic Lupus Erythematosus (CXCL10) and Positive Regulation of Th2 Cytokines (IL6) signatures. Points are colored by density as in **B**; lines represent linear model fit with 95% confidence interval. **E.** Heatmap displaying changes in circulating levels of proteins in the DNA Methylation signature that are significantly positively correlated with CRP levels. The left column represents Spearman *rho* values for correlation with CRP values, while the right columns display median Z-scores for each feature for COVID-19-negative (-) versus COVID19-positive patients (+). Z-scores were calculated from the adjusted values for each SOMAmer in each sample, based on the mean and standard deviation of COVID-19-negative samples. Asterisks indicate a significant difference between COVID-19 patients and the control group. **F. Top**, scatter plot for correlation of CRP with H2AFZ. Points are colored by density as in **B**; lines represent linear model fit with 95% confidence interval. **Bottom**, Sina plot for H2AFZ with boxes indicating median and interquartile range. **G.** Heatmap displaying changes in circulating levels of proteins in the Response to Heat group as described for **C. H.** Data for HSPA1A as described for **F**. q-values in **F** and **H** are derived from mix linear models.

In order to investigate associations between CRP levels and underlying pathophysiological processes, we first performed Metascape pathway enrichment analysis of the positively correlated proteins measured by SOMAscan®. Somewhat expectedly, this analysis revealed enrichment of several groups of proteins associated with immune activation, such as signatures associated with Systemic Lupus Erythematosus (SLE, e.g. CXCL10), Positive Regulation of Th2 Cytokines (e.g. IL6), Acute Inflammatory Response, Cytolysis (composed mostly of complement subunits), and IFN-γ-mediated Signaling Pathway (**Figure 3C-D**). Interestingly, this analysis also identified protein signatures associated with DNA Methylation and Response to Heat. The DNA methylation group is comprised of 19 features including chromatin-associated factors (e.g., DNMT3L, DPY30, SUDS3, RBBP4, RBBP5) and several histones (e.g., HIST1H3A, H2AFZ, HIST2HA, HIST2H2BB) (**Figure 3E**). For example, H2AFZ (H2AZ) is significantly correlated with CRP and elevated in the plasma of COVID-19 patients (**Figure 3F**). The Response to Heat signature has 12 features, four of which are heat shock proteins (HSPA1A, HSPA1B, HSP90AA1, HSPH1) including HSPA1A (HSP72) which is both significantly correlated with CRP and elevated in COVID-19 (**Figure 3G-H**). Interestingly, both histones and heat shock proteins can function as Damage Associated Molecular Patterns (DAMP) molecules whose presence in the bloodstream is consistently associated with tissue damage and trauma, and which in turn function as ligands for amplification of innate immune signaling (Huang et al., 2011). Circulating histones can be released from dying cells in the liver and they have been shown to drive downstream damage to both pulmonary and hepatic endothelial cells (Kawai et al., 2016). Histones in the bloodstream could also be interpreted as a sign of netosis and formation of neutrophil extracellular traps (NETS) (Papayannopoulos, 2018). HSPs have also been identified as DAMPs produced by a number of tissues upon injury, including liver (Martin-Murphy et al., 2010) and kidney (Sabapathy et al., 2020), further exacerbating inflammation at the damaged tissue.

Notably, analysis of the correlations between CRP levels and mRNAs measured in the whole blood transcriptome identifies several histone mRNAs among the top correlations (**Figure S2A**, **Supp. File 9**). In fact, the most positively correlated mRNA is H2BC12, one of the H2B-encoding genes, and the fifth most correlated gene is H2AC19, one of the H2A-encoding genes, both elevated in the whole blood transcriptome of COVID-19-positive patients (**Figure S2I**). Given that the mRNAs captured in this transcriptome analysis are derived from circulating immune cells and that histone mRNAs are transcribed during the S phase of the cell cycle, this could be interpreted as a sign of immune cell activation and proliferation (and potentially cytolysis) in patients with higher CRP levels.

Altogether, these results indicate that high CRP levels in COVID-19 associate not only with activation of inflammatory pathways, but also with elevated DAMPs, indicative of tissue damage.

### CRP levels associate with depletion of key protective serpins

Analysis of the proteins negatively correlated with CRP levels revealed that in both proteomics datasets the most anti-correlated proteins are SERPINA5 (Protein C Inhibitor, PCI, Plasminogen Activator Inhibitor 3, PAI3) and SERPINA4 (Kallistatin), two members of the serpin family of serine protease inhibitors (**Figure 4A-B, Figure S2B-C, Figure S3, Supp. Files 10-11**). Both SERPINA5 and SERPINA4 play protective roles during vascular and organ injury (Chao et al., 2016; Suzuki, 2008), but the mechanisms driving these protective effects remain to be elucidated. SERPINA5 is a multifunctional serpin that can act as both a procoagulant via inhibition of activated protein C and thrombin, but also as an anticoagulant by inhibiting several coagulation factors including plasma kallikrein (KLKB1, kallikrein B1) (Meijers et al., 1988), tissue kallikreins (Ecke et al., 1992), prothrombin, and factors XI and Xa, among others (Suzuki, 2008). SERPINA4/Kallistatin is a potent inhibitor of tissue-specific kallikreins (KLKs) (Chao et al., 2016). Notably, both of these serpins converge on inhibition of KLKs, a family of serine proteases involved not only in control of coagulation and fibrinolysis, but also production of vasoactive kinin peptides, such as bradykinin, as well as activation of the complement cascade (Irmscher et al., 2018; Ricklin and Lambris, 2007). Therefore, we investigated if CRP levels correlated significantly with dysregulation of components of the interconnected coagulation and complement cascades (**Supp. File 16**). Indeed, CRP correlated negatively with circulating levels of both the plasma kallikrein KLKB1 and the tissue kallikrein KLK13, which are depleted in COVID-19 (**Figure 4D,E**), and positively with numerous complement subunits upregulated in COVID-19 including C9, C5, C3 and C2, among others (**Figure 4C,F,G**).

**Figure 4.**
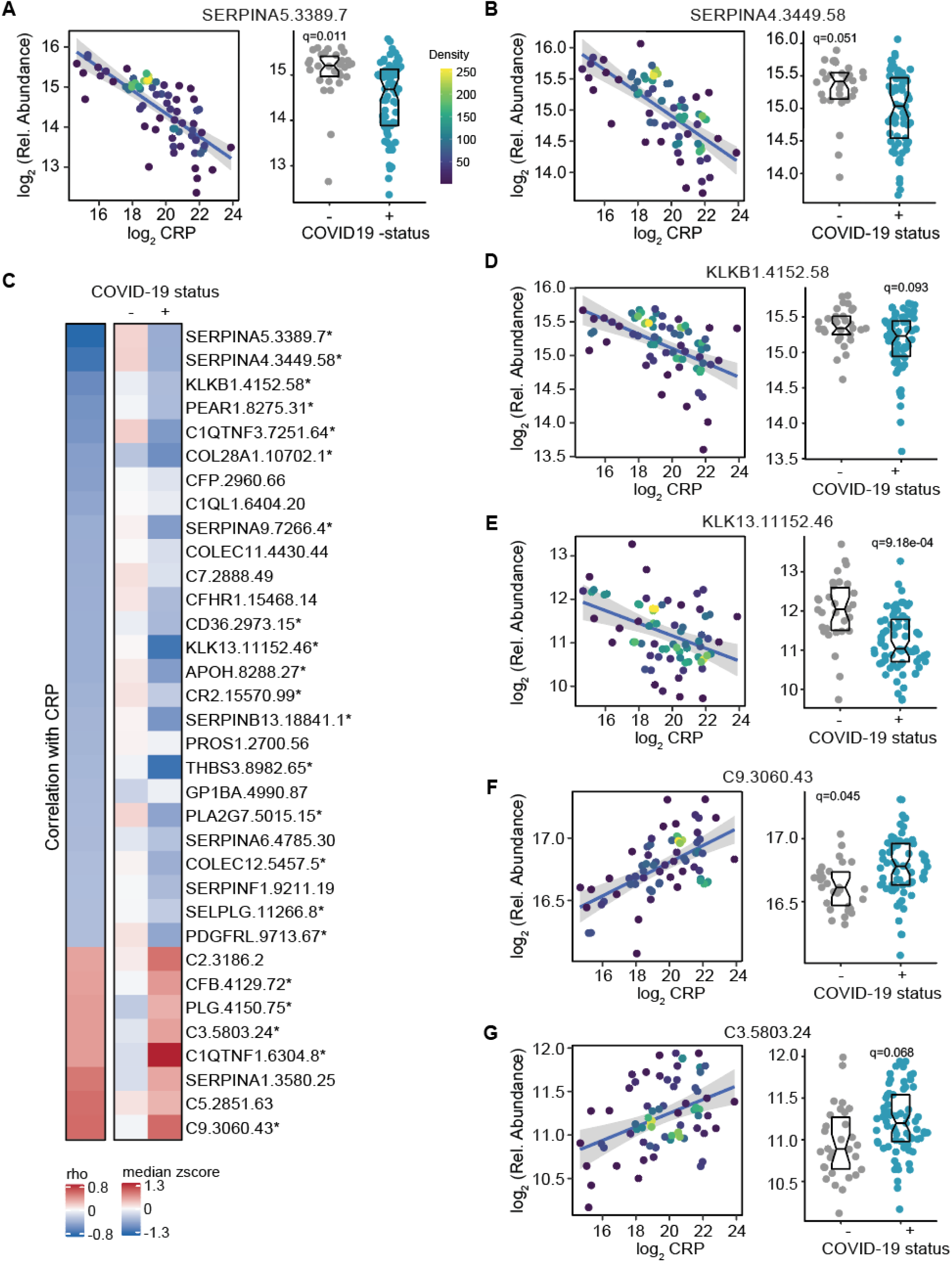
CRP levels correlate with depletion of protective serpins. **A-B.** Correlation analysis of CRP with SERPINA5 (A) and SERPINA4 (B) **Left,** scatter plot for correlation of CRP with the indicated SOMAmer® reagent. Points are colored by density; lines represent linear model fit with 95% confidence interval. **Right**, Sina plot for indicated SOMAmer® reagent with boxes indicating median and interquartile range. **C.** Heatmap displaying changes in circulating levels of complement and coagulation proteins significantly correlated with CRP levels with an absolute *rho* value greater than 0.3. The left column represents Spearman *rho* values, while the right columns display median Z-scores for each feature for COVID-19-negative controls (-) versus COVID-19-positive patients (+). Z-scores were calculated from the adjusted values for each SOMAmer® in each sample, based on the mean and standard deviation of COVID-19-negative samples. Asterisks indicate a significant difference COVID-19 patients and the control group. **D-G.** Scatter and sina plots as in **A** for KLKB1, KLK13, C9, and C3, respectively. q-values in each are derived from linear models.

Altogether, these results indicated that the prognostic value of high CRP levels in COVID-19 could be potentially tied to the accompanying depletion of important protective serpins and consequent dysregulation of the coagulation, fibrinolysis, and complement cascades, both of which have been involved in the etiology of severe COVID-19 pathology (Lo et al., 2020).

### CRP associates with dysregulated mitochondrial metabolism in peripheral blood cells

Next, we investigated associations between CRP levels and metabolic changes detected in the plasma and RBC metabolomics datasets. CRP correlated significantly with 25 plasma metabolites and 13 RBC metabolites (**Figure S2E-F, Supp. Files 13-14**). The tryptophan catabolite kynurenine is significantly positively correlated with CRP levels in both metabolomics datasets (**Figure S2E-F, Supp. Files 13-14**). Activation of the kynurenine pathway is often associated with inflammation and has been demonstrated in COVID-19 (Thomas et al., 2020). Further analysis of the top positive correlations in the plasma metabolome revealed multiple associations indicative of dysregulated mitochondrial metabolism in patients with elevated CRP. Three different carbon sources for the tricarboxylic acid (TCA) cycle were positively correlated with CRP, including the branched chain amino acids leucine (the most positively correlated metabolite) and isoleucine; pyruvate; and several acyl-carnitines (e.g. O-dodecenoyl-carnitine, tetradecenoyl carnitine, O-dodecanoyl-carnitine) (**Figure 5A-B**, **Supp. File 13**). Lactate, which can be oxidized to pyruvate by lactate dehydrogenase, was also significantly positively correlated with CRP (**Figure 5A**). Increases in lactate and pyruvate can be interpreted as increased glycolysis in patients with high CRP, perhaps driven by hypoxia of carbon flow from pyruvate to acetyl-CoA. Increased glycolysis is a metabolic consequence of both immune cell activation and hypoxemia (Frauwirth et al., 2002; Jellusova, 2020; Makowski et al., 2020; Michalek et al., 2011; van Teijlingen Bakker and Pearce, 2020). Importantly, each of these three classes of metabolites represent entry points to the TCA cycle and elevated levels of these features are consistent with mitochondrial dysfunction and decreased activity in the TCA cycle and the electron transport chain (ETC).

**Figure 5.**
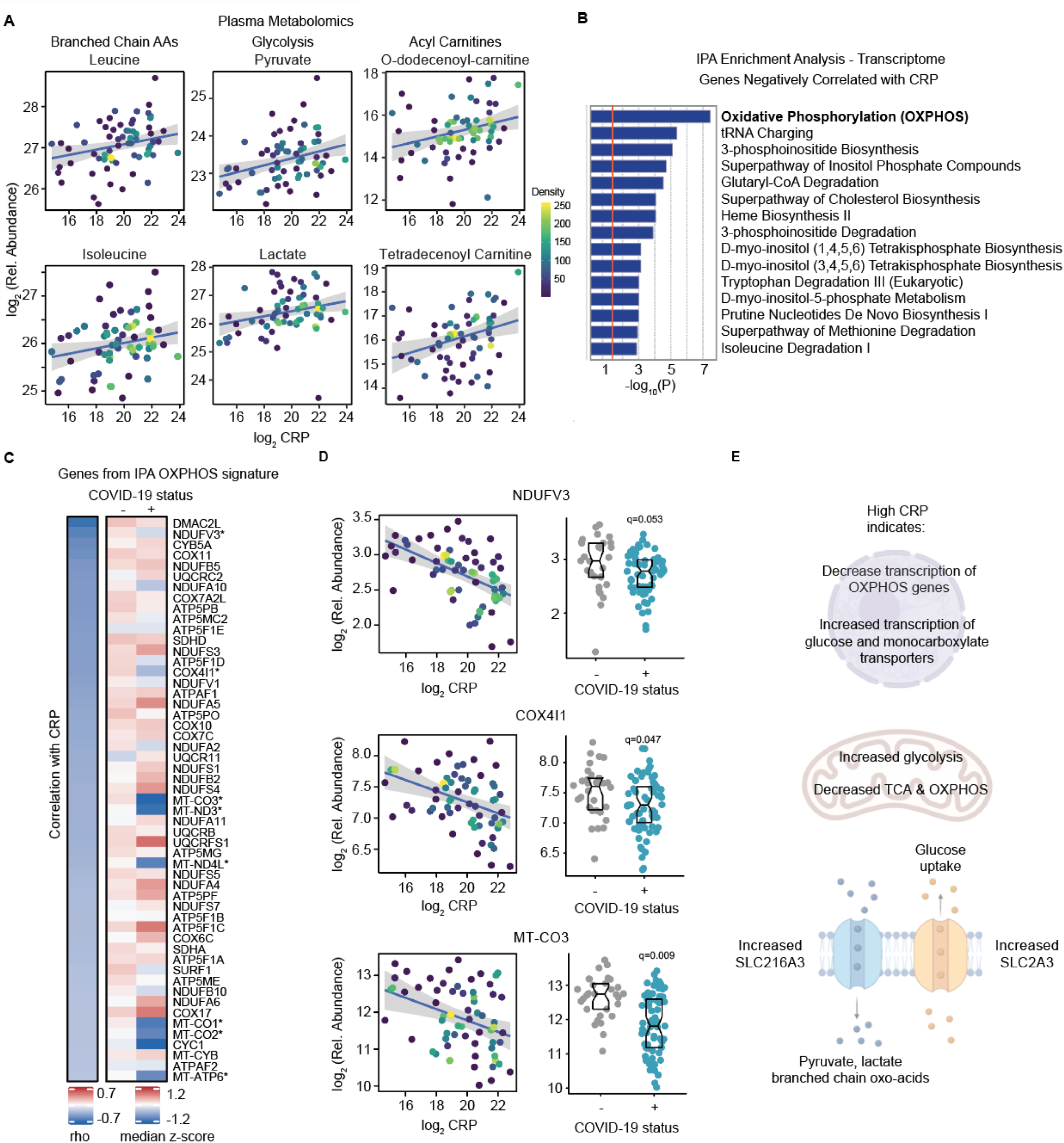
CRP levels correlate with dysregulated mitochondrial metabolism in blood cells. **A.** Scatter plot dislpaying correlations between CRP levels and indicated metabolites. Points are colored by density; lines represent linear model fit with 95% confidence interval. **B.** Histogram displaying the results of Ingenuity Pathway Analysis (IPA) of metabolic pathways for mRNAs measured in the whole blood transcriptome analysis that are significantly and negatively correlated with CRP. **C.** Heatmap displaying expression changes in mRNAs in the Oxidative Phosphorylation (OXPHOS) IPA signature from **B**. The left column represents Spearman *rho* values for correlations with CRP, while the right columns display median Z-scores for each feature for COVID-19-negative controls (-) versus COVID-19-positive patients (+). Z-scores were calculated from the adjusted RPKM values for each mRNA in each sample, based on the mean and standard deviation of COVID-19-negative samples. Asterisks indicate a significant difference between COVID-19 patients and the control group. **D. Left,** scatter plots for correlations between CRP and the indicated mRNAs. Points are colored by density as in **A**; lines represent linear model fit with 95% confidence interval. **Right**, Sina plots for indicated mRNAs with boxes indicating median and interquartile range. q-values in each sina plot are from DESeq2. **E.** Summary of findings indicating dysregulation of mitochondrial metabolism in the bloodstream of COVID-19 paients. Created with graphic elements from BioRender.com.

Given that these metabolic associations between CRP and plasma metabolites could be due to metabolic dysregulation in peripheral blood cells and/or various host tissues, we asked if these associations could be explained by gene expression changes in circulating blood cells by analyzing the whole blood transcriptome dataset. We used the Ingenuity Pathway Analysis (IPA) software to identify gene sets enriched among the RNAs positively and negatively correlated with CRP, with a focus on metabolic pathways. Strikingly, the most significantly enriched metabolic pathway among negatively correlated mRNAs is Oxidative Phosphorylation (OXPHOS) (**Figure 5B**, see **Figure S4A** for positively correlated gene sets). The Oxidative Phosphorylation gene signature is comprised of 54 genes including many components of the ETC, such as NADH:ubiquinone oxidoreductase subunits, cytochrome C complex subunits, and ATP synthase subunits, among others (**Figure 5C**). For example, expression of NDUFV3 (NADH:Ubiquinone Oxidoreductase Subunit V3, Complex I, Mitochondrial Respiratory Chain, 10-KD Subunit) and COX4I1 (Cytochrome C Oxidase Subunit 4I1) are both negatively correlated with CRP levels and significantly decreased in COVID-19 patients, as is the Mitochondrially Encoded Cytochrome C Oxidase III (MT-CO3) (**Figure 5D**). Therefore, accumulation of TCA carbon sources in plasma could be linked to decreased gene expression of ETC components in circulating blood cells. Interestingly, we noticed that the mRNAs encoding the glucose transporter SLC2A3 (GLUT3) and the monocarboxylate transporter SLC16A3 (monocarboxylate transporter 4, MCT4), were both positively and significantly correlated with CRP in the whole blood transcriptome of COVID-19 patients (**Figure S4B**). Increases in surface expression of SLC2A3 have been noted during activation of diverse lymphocytes, neutrophils and platelets, and thought to mediate increased glucose uptake to fuel cell activation (Simpson et al., 2008). SLC16A3 catalyzes the bidirectional transport across the plasma membrane of many monocarboxylates such as lactate, pyruvate, as well as branched-chain oxo-acids derived from leucine, valine and isoleucine. In innate immune cells, lactate is produced and exported in large amounts via SLC16A3 during pro-inflammatory responses and its expression is upregulated in activated macrophages (Weiss and Angiari, 2020). Furthermore, SLC16A3 expression is necessary for macrophage activation, as its deletion results in intracellular accumulation of lactate and decreased glycolysis (Weiss and Angiari, 2020).

Altogether, the metabolic changes associated with CRP could be understood, in part, as the byproduct of metabolic remodeling of circulating blood cells, whereby decreased expression of OXPHOS genes and increased expression of glucose and monocarboxylate transporters would lead to increased glucose uptake, decreased OXPHOS, and consequent accumulation of glycolysis end products (lactate, pyruvate) and other carbon sources for the TCA cycle (branched chain amino acids, acyl carnitines).

## DISCUSSION

The global health crisis imposed by the COVID-19 pandemic has inspired new approaches for rapid collaboration, open access to manuscripts under review, and data sharing. Here, we describe the rapid creation of a user-friendly researcher portal enabling easy access and real-time analysis of matched multi-omics datasets for COVID-19. The first batch of biospecimens for the COVIDome project was received by this team in July 2020, and the COVIDome Explorer was publicly launched in November 2020, thus spanning only five months from sample processing to portal launch. Between November 2020 and February 2021, more than 500 unique users had utilized the portal according to session data gathered from Google Analytics. Currently, the second batch of samples is being subjected to identical multi-omics analyses. Importantly, the COVIDome Explorer can easily ingest datasets from other teams to be displayed in its dashboards, which would then enable the comparison of results across different studies.

With the advent of multi-omics platforms, it is now possible to rapidly investigate hundreds of molecular, cellular, and physiological processes from a single biospecimen. Such a systems biology approach enables the integration of findings across different methodologies and layers of biological information to expedite the pace of discovery into the etiology of a medical condition. In this report, we illustrate the power of this approach by exploring biosignatures associated with CRP, a well characterized marker of inflammation across a numerous medical conditions, including COVID-19. Although it is well established that CRP levels and trajectory have prognostic value in COVID-19 (Mousavi-Nasab et al., 2020; Mueller et al., 2020; Sharifpour et al., 2020), the exact pathophysiological processes associated with this clinical biomarker of inflammation remain to be fully elucidated. What exactly is being revealed by high baseline levels and/or rapidly elevation of CRP in COVID-19? Our analysis demonstrates that, in addition to the well-established links between CRP and other markers of inflammation and immune activity, CRP levels associate with DAMPs, depletion of protective serpins, and dysregulation of mitochondrial metabolism in blood cells in COVID-19.

The association between CRP and DAMPs reveals that CRP levels inform about the extent of tissue damage in COVID-19. High levels of CRP associate with increased circulating levels of intracellular proteins released into the bloodstream during organ damage (e.g. histones, HSPs), and which in turn can further exacerbate the inflammatory phenotype. In turn, increased tissue damage could be conceptually tied to the clear depletion of the protective serpins SERPINA4 and SERPINA5, the most anti-correlated proteins with CRP in our proteomics datasets. SERPINA4/5 depletion could lead to exacerbated, harmful levels of activity within the coagulation system, kallikrein-kinin system, and complement cascade, all of which can contribute to COVID-19 pathology. Depletion of SERPINA5 could unleash a protease storm within the coagulation cascade, leading to coagulopathies and thromboembolism in COVID-19 (Becker, 2020). Given that both SERPINA4 and SERPINA5 inhibit KLKs, the serine proteases driving production of the vasoactive peptide bradykinin, their depletion could contribute to the so called ‘bradykinin storm’ in COVID-19 linked to accumulation of fluids in the lungs and respiratory failure (Garvin et al., 2020). Lastly, since KLKs also activate the complement cascade, SERPINA4/5 depletion could lead to harmfully high levels of complement activity and consequent tissue damage by the membrane attack complex (MAC). Of note, all these processes are suitable to pharmacological modulation and the focus of many ongoing clinical trials testing the efficacy of blood thinners (Rentsch et al., 2021), kinin receptor antagonists (van de Veerdonk et al., 2020), and complement inhibitors (Mastellos et al., 2021) in COVID-19. Therefore, we posit that CRP could serve as a biomarker to stratify the patient cohorts in these clinical trials to assess potential differences between individuals with varying CRP levels. We also hypothesize that SERPINA4 and/or SERPINA5 administration could be a valid therapeutic strategy in COVID-19 to reduce organ damage, especially in patients with high CRP levels (Rau et al., 2007; Suzuki, 2008).

Interpretation of the association between CRP levels and markers of dysregulated mitochondrial metabolism must consider a combination of metabolic effects on circulating blood cells and host tissues. Plasma metabolomics can inform about metabolic alterations in the peripheral immune cell repertoire, platelets and RBCs, but also about dysregulated metabolism in various organs. CRP levels correlated with increased levels of three different carbon sources for the TCA cycle: branched chain amino acids (leucine, isoleucine), end products of glycolysis (lactate, pyruvate), and acyl carnitines, all of which could be explained by decreased activity in the TCA cycle and electron transport chain (ETC). Indeed, when analyzing the transcriptome of circulating immune cells, the top gene signature negatively associated with CRP was Oxidative Phosphorylation, with expression of many components of the ETC being downregulated in patients with high CRP. Furthermore, these changes were accompanied by increase mRNA expression of the glucose transporter SLC2A3 and the monocarboxylate transporter SLC16A3, which can be associated to activation of different immune cell subsets (Simpson et al., 2008; Weiss and Angiari, 2020). Dysregulation of mitochondrial metabolism is increasingly appreciated in COVID-19 (Burtscher et al., 2020). Notably, disruption of the TCA cycle has been reported downstream of inflammatory stimulation via a mechanism shunting citrate to succinate driving additional inflammation, largely in myeloid cells (Makowski et al., 2020; Mills et al., 2016; Tannahill et al., 2013).

Importantly, downregulation of oxidative phosphorylation and ETC genes has been demonstrated in the liver in the case of hepatitis C infection (Gerresheim et al., 2019), and in the diaphragm, liver, and peripheral blood during sepsis (Callahan and Supinski, 2005; Eyenga et al., 2014; Weiss et al., 2014). In sum, the COVIDome datasets and the COVIDome Explorer facilitate rapid hypothesis generation and testing, revealing unexpected associations between diverse molecular, cellular, and pathophysiological processes in COVID-19, even for well-studied factors such as CRP.

## Data Availability

All data generated for this manuscript is made available through the online researcher gateway of the COVIDome Project, known as the COVIDome Explorer, which can be accessed at covidome.org. The RNAseq data have been deposited in NCBI Gene Expression Omnibus, with series accession number GSE167000. The mass spectrometry proteomics data have been deposited to the ProteomeXchange Consortium via the PRIDE partner repository (Perez-Riverol et al., 2019) with the dataset identifier PXD022817. The mass cytometry data has been deposited in Flow Repository under the link: https://flowrepository.org/id/RvFrSYioKeUdYHXdkTD9TQPAXt4PqdkB5eie82h11JgAGSCQIneLKpcKd81Nzgwq. The SOMAscan Proteomics, MSD Cytokine Profiles, and Sample Metadata files have been deposited in Mendeley under entry doi:10.17632/2mc6rrc5j3.1. The metabolomics data have been deposited in the Metabolomics Workbench with a Study ID to be made available upon acceptance. All code required to run the COVIDome Explorer applications can be found at https://github.com/cusom/CUSOM.COVIDome.Shiny-Apps and https://github.com/cusom/CUSOM.ShinyHelpers.

https://github.com/cusom/CUSOM.COVIDome.Shiny-Apps

https://github.com/cusom/CUSOM.ShinyHelpers

https://data.mendeley.com/datasets/2mc6rrc5j3/1

http://proteomecentral.proteomexchange.org/cgi/GetDataset?ID=PXD022817

https://www.ncbi.nlm.nih.gov/geo/query/acc.cgi?acc=GSE167000

## ACKNOWLEDGMENTS

We are grateful to Dr. Thomas Flaig and the Office of the Vice Chancellor For Research at the University of Colorado Anschutz Medical Campus for their leadership in setting up the COVID-19 Biobank at the University of Colorado and also to the COVID-19 Biobank Steering Committee for overall support of this project. We thank members of the Biorepository Shared Resource, especially Dr. Adrie Van Bokhoven, Zachary Grasmick, and Hannah Schumman; members of the Human Immune Monitoring Shared Resource, especially Dr. Jill Slansky, Jodi Livesay, Troy Schedin, and Dr. Jennifer McWilliams; members of the Flow Cytometry Shared Resource of the University of Colorado Cancer Center, specially Dr. Eric Clambey, Alistair Acosta, Christine Childs, and Kristina Terrell; as well as Dr. Aaron Issaian for assistance with MS proteomics data analysis. We also thank the SomaLogic team for their support and the Meso Scale Discovery team for generous support with seroconversion assays. We are grateful to Dr. Ian Brooks, Dr. Michelle Edelman and the Health Data Compass Data Warehouse project (healthdatacompass.org) for the clinical data.

## AUTHOR CONTRIBUTIONS

JME designed the project and organized the multiple collaborations required for creation of the COVIDome dataset and the COVIDome Explorer. KB, NL, and MGM built the COVIDome database and Researcher Portal. KDS, MDG, KTK, PA, KPS, REG, RB, KRJ, SR, MD, JAR, RC, TC, AAM, TDB, EWYH, AD, KCH, and JME, designed experiments and analyzed data. KDS and JME wrote the manuscript. All authors reviewed the manuscript.

## DECLARATION OF INTERESTS

KDS and JME are co-inventors on two patents related to JAK inhibition in COVID-19; JME serves in the COVID Development Advisory Board for Elly Lilly and has provided consulting services to Gilead Sciences Inc. JME serves on the Cell Reports Advisory Board.

**Supplementary File 1. Cohort Characteristics.** Table summarizing cohort characteristics. Information pertaining less than 10% of the cohort is indicated as <10% to prevent potential reidentification. Information pertaining to less than 10 participants is indicated as <10 to prevent potential reidentification. For clinical labs, values represent the mean -/+ standard deviation from the mean. Acronyms for clinical laboratory measurements are as follows: BUN: blood urea nitrogen; CRP: C-reactive protein; ALT: alanine aminotransferase; ALP: alkaline phosphatase; AST: aspartate aminotransferase; BNP: brain natriuretic peptide. Comorbidities affecting each group are listed based on two different annotations: Carlson and Elixhauser. Acronyms for comorbidities are as follows: CHF: chronic heart failure; DM: diabetes mellitus; DMCX: diabetes with complications; METS: metastatic cancer; MI: myocardial infarction; PUD: peptic ulcer disease; PVD: peripheral vascular disease; HTN: hypertension; PHTN: pulmonary hypertension. Fisher’s exact test was used to calculate p values for differences in % among groups, and the Mann-Whitney test was used to calculate p values for differences in clinical lab values.

**Supplementary File 2. Transcriptome.** Differential expression analysis of COVID-19-positive versus COVID-19-negative patients using DESeq2. Columns include: (**A**) gene name, (**B**) chromosome, (**C**) Ensemble gene ID, (**D**) baseMean of all samples, (**E**) baseMean of COVID-19-negative samples, (**F**) baseMean of COVID-19-positive samples, (**G**) adjusted fold change, (**H**) adjusted log_2_ fold change, (**I**) p-value, (**J**) adjusted p-value, (**K**) gene start coordinate, (**L**) gene end coordinate, (**M**) gene type, and (**N**) HGNC ID.

**Supplementary File 3. SOMAscan® Proteomics.** Differential abundance analysis of SOMAscan® proteomics from COVID-19-positive versus COVID-19-negative patients using a linear model. Columns include (**A**) aptamer name, (**B**) analyte, (**C**) analyte description, (**D**) Entrez gene symbol, (**E**) Entrez gene ID, (**F**) Average value of COVID-19-negative samples, (**G**) Average value of COVID-19-positive samples, (**H**) fold change, (**I**) log_2_ fold change, (**J**) p-value, and (**K**) adjusted p-value (q-value) via Bonferroni-Hochberg (BH) method.

**Supplementary File 4. Mass Spectrometry Plasma Proteomics.** Differential abundance analysis of MS proteomics from COVID-19-positive versus COVID-19-negative patients using a linear model adjusting for age and sex. Columns include (**A**) analyte, (**B**) analyte description, (**C**) SwissProt ID, (**D**) average value of COVID-19-negative samples, (**E**) average value of COVID-19-positive samples, (**F**) fold change, (**G**) log_2_ fold change, (**H**) p-value, and (**I**) adjusted p-value (q-value) via Bonferroni-Hochberg (BH) method.

**Supplementary File 5. Meso Scale Discovery (MSD) Cytokine Profiling.** Differential abundance analysis of cytokines from COVID-19-positive versus COVID-19-negative patients using a linear model adjusting for age and sex. Columns include (**A**) Analyte, (**B**) average value of COVID-19-negative samples, (**C**) average value of COVID-19-positive samples, (**D**) fold change, (**E**) log_2_ fold change, (**F**) p-value, and (**G**) adjusted p-value (q-value) via Bonferroni-Hochberg (BH) method.

**Supplementary File 6. Red Blood Cell (RBC) Metabolomics.** Differential abundance analysis of MS RBC Metabolomics from COVID-19-positive versus COVID-19-negative patients using a linear model adjusting for age and sex. Columns include (**A**) analyte, (**B**) average value of COVID-19-negative samples, (**C**) average value of COVID-19-positive samples, (**D**) fold change, (**E**) log_2_ fold change, (**F**) p-value, and (**G**) adjusted p-value (q-value) via Bonferroni-Hochberg (BH) method.

**Supplementary File 7. Plasma Metabolomics.** Differential abundance analysis of MS plasma Metabolomics from COVID-19-positive versus COVID-19-negative patients using a linear model. Columns include **A**) analyte, (**B**) average value of COVID-19-negative samples, (**C**) average value of COVID-19-positive samples, (**D**) fold change, (**E**) log_2_ fold change, (**F**) p-value, and (**G**) adjusted p-value (q-value) via Bonferroni-Hochberg (BH) method.

**Supplementary File 8. Mass Cytometry.** Differential abundance analysis of immune cell types from COVID-19-positive versus COVID-19-negative patients using a linear model. Columns include (**A**) population, (**B**) definition of population, (**C**) average value of COVID-19-negative samples, (**D**) average value of COVID-19-positive samples, (**E**) fold change, (**F**) log_2_ fold change, (**G**) p-value, and (**H**) adjusted p-value using Benjamini-Hochberg method. Tabs include analysis of all live cells, CD3+ T cells, CD4+ T cells, CD8+ T cells, CD19+ B cells, Monocytes, and Myeloid DCs.

**Supplementary File 9. CRP-Transcriptome Correlations.** Results of Spearman correlation analysis between mass spectrometry CRP levels and transcripts detected by whole blood RNAseq analysis. Columns include: (**A**) Ensembl gene ID, (**B**) gene name, (**C**) Spearman *rho* value, (**D**) p-value, and (**E**) adjusted p-value (q-value) via Bonferroni-Hochberg (BH) method.

**Supplementary File 10. CRP-MS Plasma Proteomics Correlations.** Results of Spearman correlation analysis between mass spectrometry CRP levels and proteins identified by mass spectrometry. Columns include: (**A**) protein ID, (**B**) SwissProt ID, (**C**) Spearman *rho* value, (**D**) p-value, and (**E**) adjusted p-value (q-value) via Bonferroni-Hochberg (BH) method.

**Supplementary File 11. CRP-SOMAscan®Proteomics Correlations.** Results of Spearman correlation analysis between mass spectrometry CRP levels and proteins identified by SOMAscan^®^ technology. Columns include: (**A**) aptamer name, (**B**) analyte, (**C**) SwissProt ID, (**D**) Gene symbol, (**E**) Spearman *rho* value, (**F**) p-value, and (**G**) adjusted p-value (q-value) via Bonferroni-Hochberg (BH) method.

**Supplementary File 12. CRP-MSD Cytokine Correlations.** Results of Spearman correlation analysis between mass spectrometry CRP levels and cytokine, chemokines, and immune factors identified by Meso Scale Discovery technology. Columns include: (**A**) analyte, (**B**) Spearman *rho* value, (**C**) p-value, and (**D**) adjusted p-value (q-value) via Bonferroni-Hochberg (BH) method.

**Supplementary File 13. CRP-Plasma Metabolomics Correlations.** Results of Spearman correlation analysis between mass spectrometry CRP levels and plasma metabolites. Columns include: (**A**) analyte, (**B**) Spearman *rho* value, (**C**) pvalue, and (**D**) adjusted p-value (q-value) via Bonferroni-Hochberg (BH) method.

**Supplementary File 14. CRP-RBC Metabolomics Correlations.** Results of Spearman correlation analysis between mass spectrometry CRP levels and red blood cell metabolites. Columns include: (**A**) analyte, (**B**) Spearman *rho* value, (**C**) p value, and (**D**) adjusted p-value (q-value) via Bonferroni-Hochberg (BH) method.

**Supplementary File 15. CRP-Mass Cytometry Correlations.** Results of Spearman correlation analysis between mass spectrometry CRP levels and immune cell populations detected by Mass Cytometry. Columns include: (**A**) population, (**B**) lineage, (**C**) Spearman *rho* value, (**D**) p value, and (**E**) adjusted p-value (q-value) via Bonferroni-Hochberg (BH) method. Tabs include analysis of all live cells, CD3+ T cells, CD4+ T cells, CD8+ T cells, CD19+ B cells, Monocytes, and Myeloid DCs.

**Supplementary File 16. CRP-Complement-Coagulation SOMAscan.** Results of Spearman correlation analysis between mass spectrometry CRP levels and complement and coagulation proteins identified by SOMAscan^®^ technology. (**A**) Aptamer name, (**B**) Spearman *rho* value, (**C**) p value, and (**D**) adjusted p-value (q-value) via Bonferroni-Hochberg (BH) method.

**Supplementary File 17. Mass Cytometry Antibody Table.** List of antibodies used in mass cytometry. Columns include: (**A**) antibody target, (**B**) element conjugated to antibody, (**C**) mass of element, (**D**) manufacturer, (**E**) catalog number, (**F**) clone number, and (**G**) staining protocol (fixed, live or fixed with permeabilization).

## STAR METHODS

### Key Resources Table

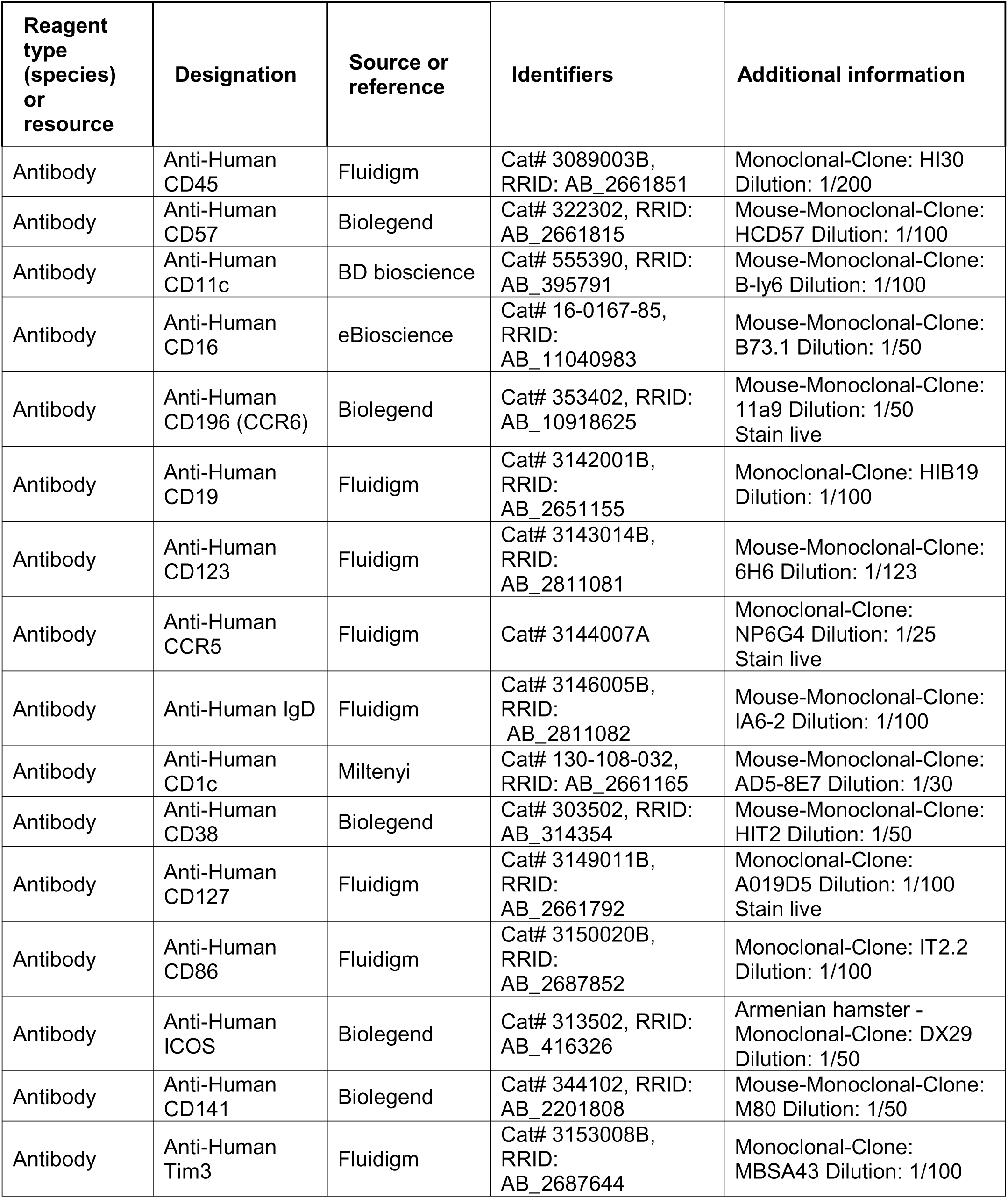
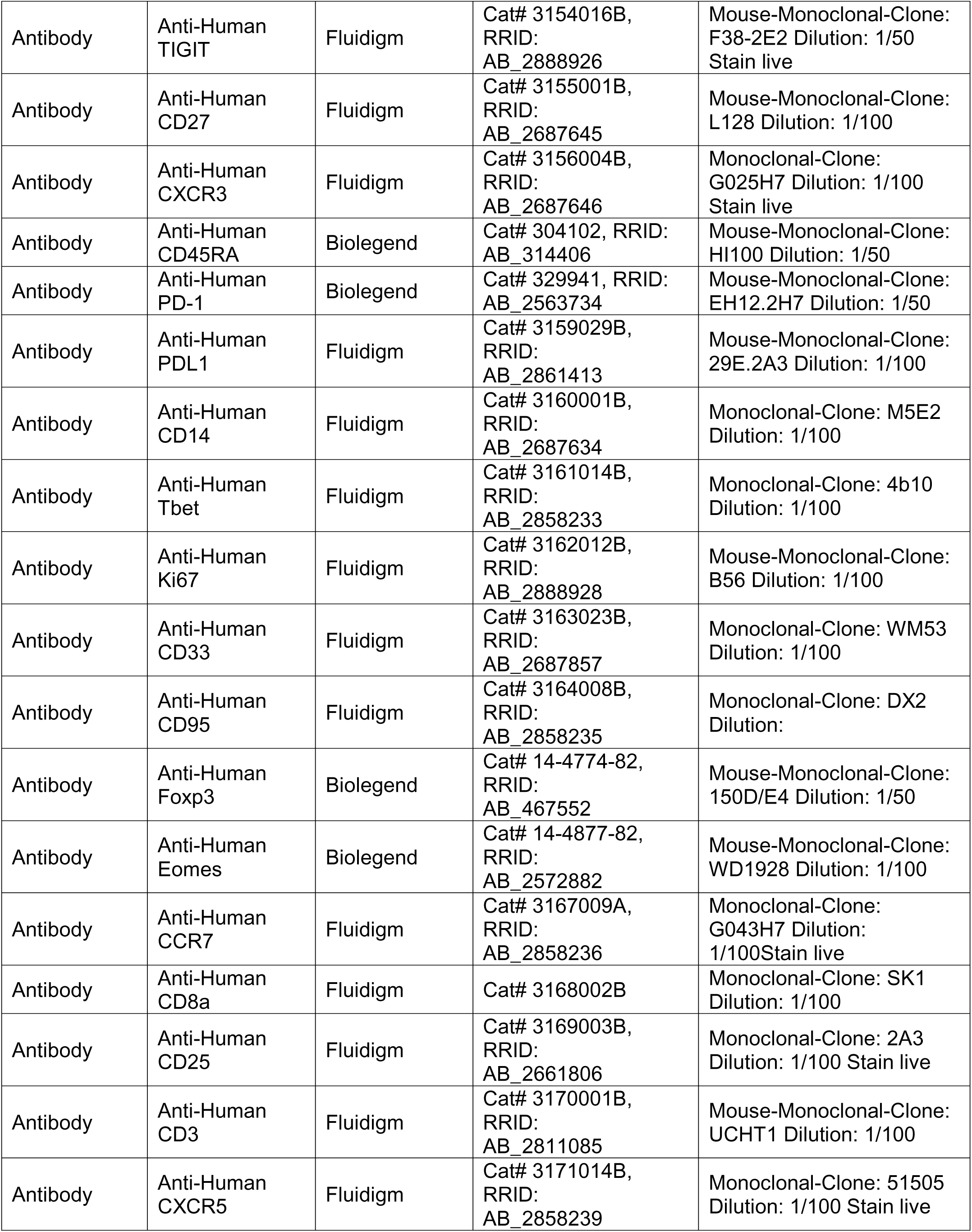
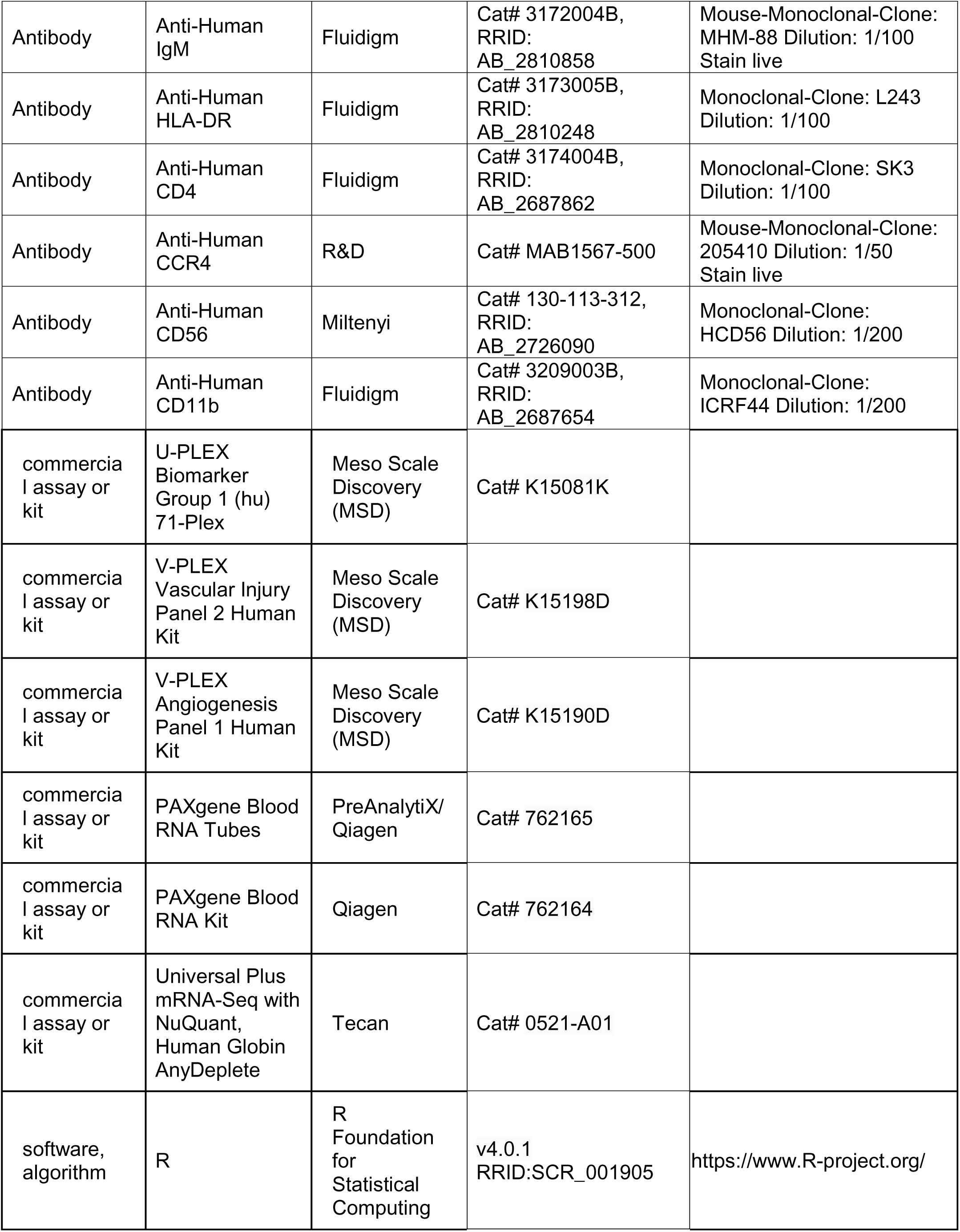
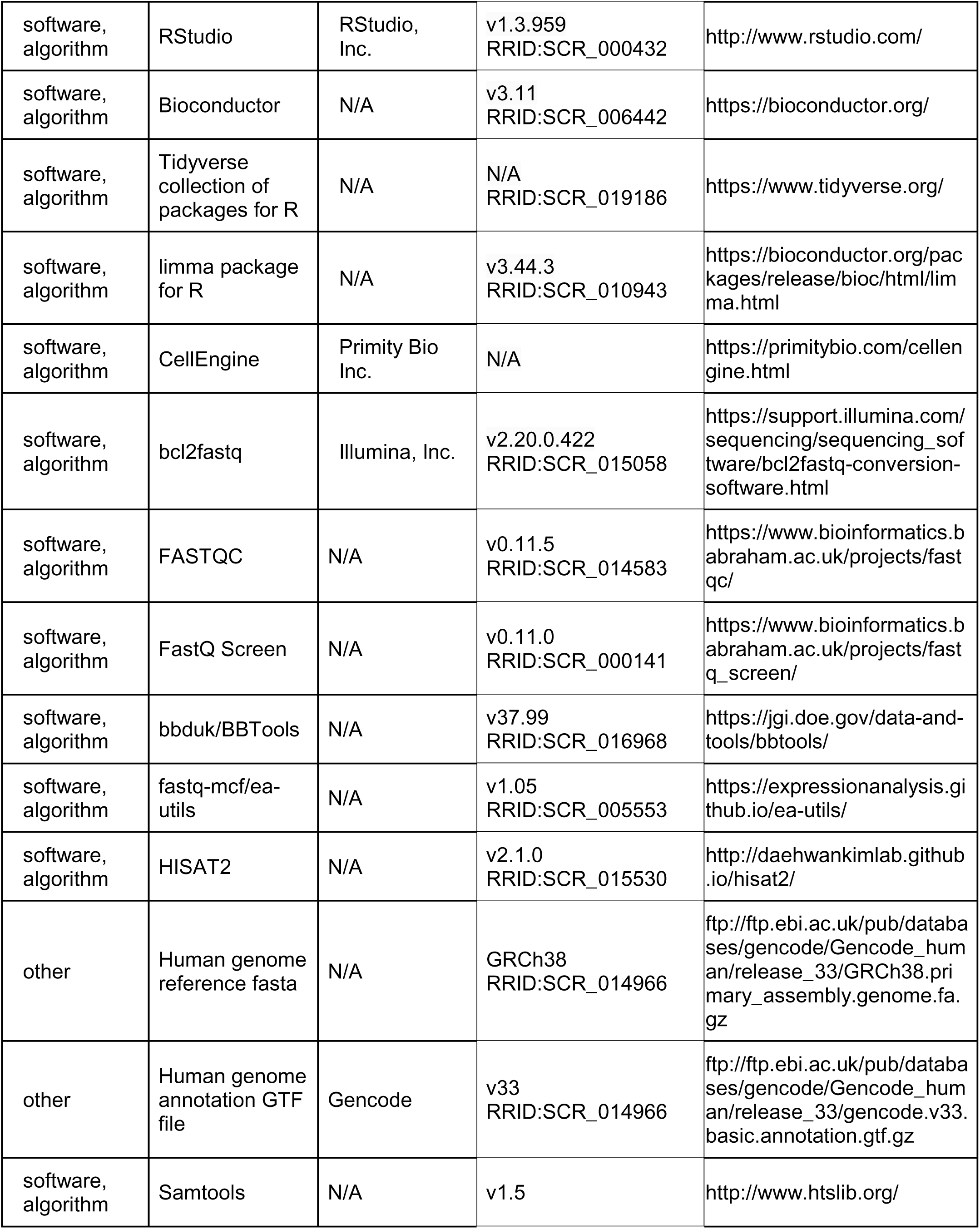
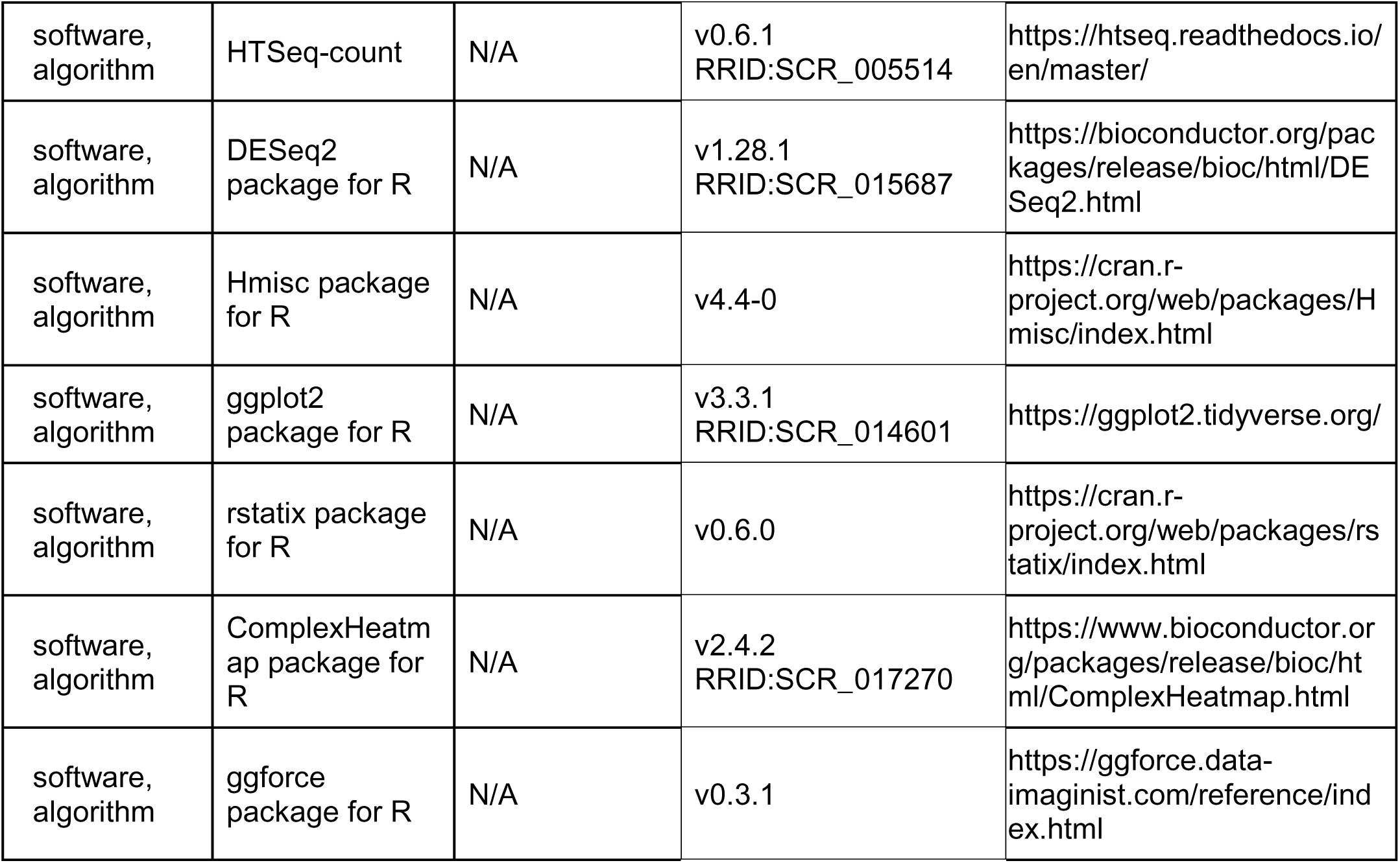

## RESOURCE AVAILABILITY

### Lead Contact

Further information and requests for resources and reagents should be directed and will be fulfilled by the Lead Contact, Joaquin Espinosa.

### Materials Availability

This study did not generate new unique reagents.

### Data and Code Availability

All data generated for this manuscript is made available through the online researcher gateway of the COVIDome Project, known as the COVIDome Explorer, which can be accessed at covidome.org. The RNAseq data have been deposited in NCBI Gene Expression Omnibus, with series accession number GSE167000. The mass spectrometry proteomics data have been deposited to the ProteomeXchange Consortium via the PRIDE partner repository (Perez-Riverol et al., 2019) with the dataset identifier PXD022817. The mass cytometry data has been deposited in Flow Repository under the link: https://flowrepository.org/id/RvFrSYioKeUdYHXdkTD9TQPAXt4PqdkB5eie82h11JgAGSCQIneLKpcKd81Nzgwq. The SOMAscan® Proteomics, MSD Cytokine Profiles, and Sample Metadata files have been deposited in Mendeley under entry doi:10.17632/2mc6rrc5j3.1. The metabolomics data have been deposited in the Metabolomics Workbench with a Study ID to be made available upon acceptance. All code required to run the COVIDome Explorer applications can be found at https://github.com/cusom/CUSOM.COVIDome.Shiny-Apps and https://github.com/cusom/CUSOM.ShinyHelpers.

## EXPERIMENTAL MODEL AND SUBJECT DETAILS

### Study design, participant recruitment, and clinical data capture

Research participants were recruited and consented for participation in the COVID Biobank of the University of Colorado Anschutz Medical Campus [Colorado Multiple Institutional Review Board (COMIRB) Protocol # 20-0685]. Data was generated from deidentified biospecimens and linked to demographics and clinical metadata procured through the Health Data Compass of the University of Colorado under COMIRB Protocol # 20-1700. Participants were hospitalized either at Children’s Hospital Colorado or the University of Colorado Hospital. COVID-19 status was defined by a positive PCR reaction and/or antibody test. Cohort characteristics can be found in **Supp. File 1**.

## METHOD DETAILS

### Blood processing

Blood samples were collected into EDTA tubes, PAXgene RNA, and sodium heparin tubes. After centrifugation, EDTA plasma was used for MS proteomics, SOMAscan^®^ proteomics, as well as multiplex immunoassays using MSD technology for both cytokine profiles and seroconversion assays. From sodium heparin tubes, PBMCs were obtained by the Ficoll gradient method before cryopreservation and assembly of batches for MC analysis (see below).

### Whole blood transcriptome

RNA was purified from PAXgene Blood RNA Tubes (PreAnalytiX/Qiagen) using a PAXgene Blood RNA Kit (Qiagen), according to the manufacturer’s instructions. RNA quality was assessed using an Agilent 2200 TapeStation and quantified by Qubit (Life Technologies). Globin RNA depletion, poly-A(+) RNA enrichment, and strand-specific library preparation were carried out using a Universal Plus mRNA-Seq with NuQuant, Human Globin AnyDeplete (Tecan). Paired-end 150 bp sequencing was carried out on an Illumina NovaSeq 6000 instrument by the Genomics Shared Resource at the University of Colorado Anschutz Medical Campus.

### Plasma proteomics by mass spectrometry

Plasma samples were digested in S-Trap filters (Protifi, Huntington, NY) according to the manufacturer’s procedure. Briefly, a dried protein pellet prepared from organic extraction of patient plasma was solubilized in 400 µl of 5% (w/v) SDS. Samples were reduced with 10 mM DTT at 55°C for 30 min, cooled to room temperature, and then alkylated with 25 mM iodoacetamide in the dark for 30 min. Next, a final concentration of 1.2% phosphoric acid and then six volumes of binding buffer [90% methanol; 100 mM triethylammonium bicarbonate (TEAB); pH 7.1] were added to each sample. After gentle mixing, the protein solution was loaded into an S-Trap filter, spun at 2000 rpm for 1 min, and the flow-through collected and reloaded onto the filter. This step was repeated three times, and then the filter was washed with 200 μL of binding buffer 3 times. Finally, 1 μg of sequencing-grade trypsin (Promega) and 150 μL of digestion buffer (50 mM TEAB) were added onto the filter and digestion carried out at 47 °C for 1 h. To elute peptides, three stepwise buffers were applied, 200 μL of each with one more repeat, including 50 mM TEAB, 0.2% formic acid in H_2_O, and 50% acetonitrile and 0.2% formic acid in H_2_O. The peptide solutions were pooled, lyophilized and resuspended in 1 mL of 0.1 % FA. 20 µl of each sample was loaded onto individual Evotips for desalting and then washed with 20 μL 0.1% FA followed by the addition of 100 μL storage solvent (0.1% FA) to keep the Evotips wet until analysis. The Evosep One system (Evosep, Odense, Denmark) was used to separate peptides on a Pepsep column, (150 µm internal diameter, 15 cm) packed with ReproSil C18 1.9 µm, 120A resin. The system was coupled to a timsTOF Pro mass spectrometer (Bruker Daltonics, Bremen, Germany) via a nano-electrospray ion source (Captive Spray, Bruker Daltonics). The mass spectrometer was operated in PASEF mode. The ramp time was set to 100 ms and 10 PASEF MS/MS scans per topN acquisition cycle were acquired. MS and MS/MS spectra were recorded from *m/z* 100 to 1700. The ion mobility was scanned from 0.7 to 1.50 Vs/cm^2^. Precursors for data-dependent acquisition were isolated within□±□1□Th and fragmented with an ion mobility-dependent collision energy, which was linearly increased from 20 to 59LeV in positive mode. Low-abundance precursor ions with an intensity above a threshold of 500 counts but below a target value of 20000 counts were repeatedly scheduled and otherwise dynamically excluded for 0.4 min. Raw data file conversion to peak lists in the MGF format, downstream identification, validation, filtering and quantification were managed using FragPipe version 13.0. MSFragger version 3.0 was used for database searches against a Human isoform-containing UniProt fasta file (version 08/11/2020) with decoys and common contaminants added. The identification settings were as follows: Trypsin, Specific, with a maximum of 2 missed cleavages, up to 2 isotope errors in precursor selection allowed for, 10.0 ppm as MS1 and 20.0 ppm as MS2 tolerances; fixed modifications: Carbamidomethylation of C (+57.021464 Da), variable modifications: Oxidation of M (+15.994915 Da), Acetylation of protein N-term (+42.010565 Da), Pyrolidone from peptide N-term Q or C (−17.026549 Da). The Philosopher toolkit version 3.2.9 (build 1593192429) was used for filtering of results at the peptide and protein level at 0.01 FDR. Label-free quantification was performed by AUC integration with matching between all runs using IonQuant.

### Plasma proteomics by SOMAscan^®^ assays

125 μL EDTA plasma was analyzed by SOMAscan^®^ assays using previously established protocols (Gold et al., 2012). Briefly, each of the 5000+ SOMAmer reagents binds a target peptide and is quantified on a custom Agilent hybridization chip. Normalization and calibration were performed according to SOMAscan^®^ Data Standardization and File Specification Technical Note (SSM-020) (Gold et al., 2012). The output of the SOMAscan^®^ assay is reported in relative fluorescent units (RFU).

### Cytokine profiling and seroconversion by multiplex immunoassay

Multiplex immunoassays MSD assays were performed on EDTA plasma aliquots following manufacturer’s instructions (Meso Scale Discovery, MSD). A list of immune factors measured by MSD can be found in **Supp. File 5.** Values were extrapolated against a standard curve using provided calibrators. Seroconversion assays against SARS-CoV-2 proteins and the control protein from the Flu A Hong Kong H3 virus were performed in a multiplex immunoassay using the IgG detection readout according to manufacturer’s instructions (MSD). Relative values were extrapolated against a standardized curve consisting of pooled COVID-19 positive reference plasma (Johnson et al., 2020).

### Mass cytometry analysis of immune cell types

Cryopreserved PBMCs were thawed, washed twice with Cell Staining Buffer (CSB) (Fluidigm), and counted with an automated cell counter (Countess II - Thermo Fisher Scientific). Extracellular staining on live cells was done in CSB for 30 min at room temperature, in 3-5^10^6^ cells per sample. Cells were washed with 1X PBS (Fluidigm) and stained with 1 mL of 0.25 mM cisplatin (Fluidigm) for 1 min at room temperature for exclusion of dead cells. Samples were then washed with CSB and incubated with 1.6% PFA (Electron Microscopy Sciences) during 10 min at room temperature. Samples were washed with CBS and barcoded using a Cell-IDTM 20-Plex Pd Barcoding Kit (Fluidigm) of lanthanide-tagged cell reactive metal chelators that will covalently label samples with a unique combination of palladium isotopes, then combined. Surface staining with antibodies that work on fixed epitopes was performed in CSB for 30 min at room temperature (see **Supp. File 17** for antibody information). Cells were washed twice with CSB and fixed in Fix/Perm buffer (eBioscience) for 30 min, washed in permeabilization buffer (eBioscience) twice, then intracellular factors were stained in permeabilization buffer for 45 min at 4°C. Cells were washed twice with Fix/Perm Buffer and were labeled overnight at 4°C with Cell-ID Intercalator-Ir (Fluidigm) for DNA staining. Cells were then analyzed on a Helios instrument (Fluidigm). To make all samples comparable, pre-processing of mass cytometry data included normalization within and between batches via polystyrene beads embedded with lanthanides as previously described (Finck et al., 2013). Files were debarcoded using the Matlab DebarcoderTool (Zunder et al., 2015). Then normalization again between batches relative to a reference batch based on technical replicates (Schuyler et al., 2019). Gating was performed using CellEngine (Primitybio) as previously described (Galbraith et al., 2020).

### Mass Spectrometry based metabolomics of plasma and red blood cells

#### Sample extraction

Samples were thawed on ice and extracted via a modified Folch method (chloroform/methanol/water 8:4:3), which completely inactivates other coronaviruses, such as MERS-CoV. Briefly, 20 μL of sample was diluted in 130 μL of LC-MS grade water, 600 μL of ice-cold chloroform/methanol (2:1) was added, and the samples were vortexed for 10 seconds. Samples were then incubated at 4°C for 5 minutes, quickly vortexed (5 seconds), and centrifuged at 14,000 *g* for 10 minutes at 4°C. The top (i.e., aqueous) phase was transferred to a new tube for metabolomics analysis and flash frozen. The bottom (i.e., organic) phase was transferred to a new tube for lipidomics analysis, then dried under N_2_ flow.

#### UHPLC-MS metabolomics

Analyses were performed using a Vanquish UHPLC coupled online to a Q Exactive high resolution mass spectrometer (Thermo Fisher Scientific, Bremen, Germany). Samples (10 uL per injection) were randomized and analyzed in positive and negative electrospray ionization modes (separate runs) using a 5-minute C18 gradient on a Kinetex C18 column (Phenomenex) as described (Nemkov et al., 2019). Data were analyzed using Maven (Princeton University, Princeton, NJ, USA) in conjunction with the KEGG database and an in-house standard library.

## QUANTIFICATION AND STATISTICAL ANALYSIS

Preprocessing, statistical analysis, and plot generation for all datasets was carried out using R (R 4.0.1 / Rstudio 1.3.959 / Bioconductor v 3.11) (Huber et al., 2015; R Core Team, 2020; RStudio Team, 2020), as detailed below.

### Analysis of transcriptome data

RNA-seq data yield was ∼40-80 x 10^6^ raw reads and ∼32-71 x 10^6^ final mapped reads per sample. Reads were demultiplexed and converted to fastq format using bcl2fastq (bcl2fastq v2.20.0.422). Data quality was assessed using FASTQC (v0.11.5) (https://www.bioinformatics.babraham.ac.uk/projects/fastqc/) and FastQ Screen (v0.11.0, https://www.bioinformatics.babraham.ac.uk/projects/fastq_screen/). Trimming and filtering of low-quality reads was performed using bbduk from BBTools (v37.99)(Bushnell et al., 2017) and fastq-mcf from ea-utils (v1.05, https://expressionanalysis.github.io/ea-utils/). Alignment to the human reference genome (GRCh38) was carried out using HISAT2 (v2.1.0)(Kim et al., 2019) in paired, spliced-alignment mode with a GRCh38 index with a Gencode v33 annotation GTF, and alignments were sorted and filtered for mapping quality (MAPQ > 10) using Samtools (v1.5)(Li et al., 2009). Gene-level count data were quantified using HTSeq-count (v0.6.1)(Anders et al., 2015) with the following options (--stranded=reverse –minaqual=10 –type=exon --mode=intersection-nonempty) using a Gencode v33 GTF annotation file. Differential gene expression in COVID+ versus COVID-was evaluated using DESeq2 (version 1.28.1)(Love et al., 2014) in R (version 4.0.1), using q < 0.1 (FDR < 10%) as the threshold for differentially expressed genes.

### Analysis of MS-proteomic data

Raw Razor intensity data were filtered for high abundance proteins by removing those with >70% zero values in both COVID-19-negative and COVID-19-positive groups. For the remaining 407 abundant proteins, 0 values (8,363 missing values of 44,363 total measurements) were replaced with a random value sampled from between 0 and 0.5x the minimum non-zero intensity value for that protein. Data was then normalized using a scaling factor derived from the global median intensity value across all proteins / sample median intensity across all proteins (De Livera et al., 2012) *SOMAscan^®^ data.* Normalized data (RFU) was imported and converted from a SOMAscan^®^ .adat file using a custom R package (SomaDataIO) for use in all subsequent analysis.

### Analysis of MSD cytokine profiling data

Plasma concentration values (pg/mL) for each of the cytokines and related immune factors measured across multiple MSD assay plates was imported to R, combined, and analytes with >10% of values outside of detection or fit curve range flagged. For each analyte, missing values were replaced with either the minimum (if below fit curve range) or maximum (if above fit curve range) calculated concentration and means of duplicate wells used in all further analysis.

### Analysis of LCMS-metabolomics data

Peak intensity data was imported to R. Across the 171 metabolites, 0 values (486 missing values of 21,033 total measurements) were replaced with a random value sampled from between 0 and 0.5x the minimum non-zero intensity value for that metabolite. Data was then normalized using a scaling factor derived from the global median intensity value across all proteins / sample median intensity and used for downstream analysis.

### Analysis of mass cytometry data

Cell population frequencies were exported from CellEngine as percentages of various parental lineages and used for subsequent analysis.

### Differential abundance analysis

Differential abundance analysis for MS proteomics, SOMAscan^®^ proteomics, MSD cytokine profiling, LCMS metabolomics, and CyTOF mass cytometry data was performed using linear models with log_2_ concentration as the outcome variable and age, sex, and COVID-19 status as independent variables. Multiple hypothesis correction was performed with the Benjamini-Hochberg method using a false discovery rate (FDR) threshold of 10% (q<0.1).

### Correlation analysis

To identify features in each dataset that correlate with CRP levels in COVID-19-positive samples, Spearman *rho* values and p-values were calculated against values adjusted for Sex and Age using the *removeBatchEffect* function from the limma package (v 3.44.3) (43) from each dataset using the *rcorr* function from the Hmisc package (v 4.4-0) (Jr and with contributions from Charles Dupont and many others, 2020), with Benjamini-Hochberg correction of p-values and an estimated false discovery fate threshold of 0.1. For visualization, XY scatter plots with points colored by local density were generated using a custom density function and the ggplot2 (v3.3.1) package (Wickham, 2016).

### Data Visualization

To visualize differences between COVID-19-negative samples and COVID-19-positive samples, Z-scores were calculated for each feature based on the mean and standard deviation of COVID-19-negative samples, and visualized as heatmaps and/or modified sina plots using the ComplexHeatmap (v2.4.2) (Gu et al., 2016), ggplot2 (v3.3.1), and ggforce (v0.3.1) packages (Pedersen, 2019).

**Supplementary Figure 1, related to Figure 1.**
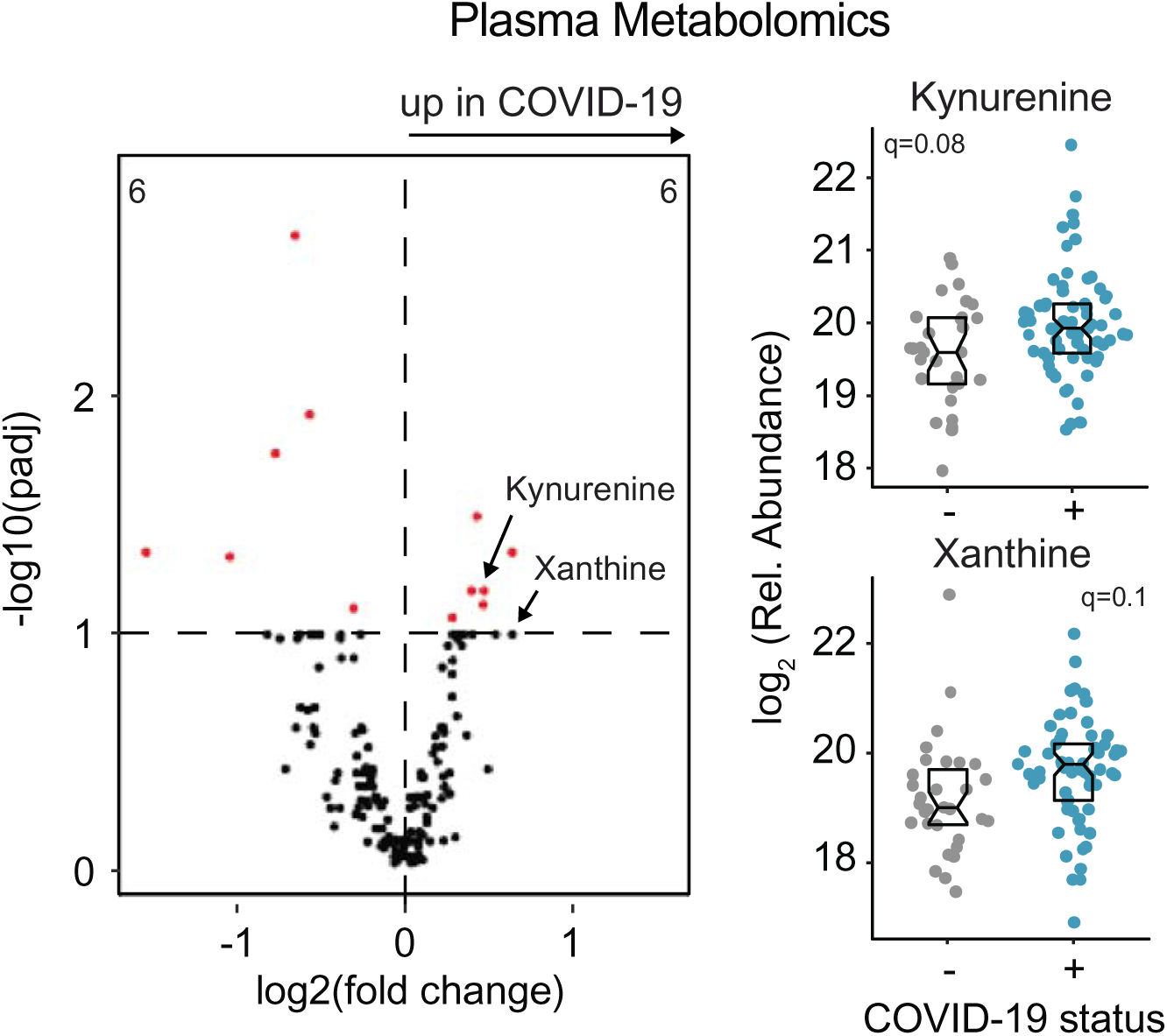
COVIDome plasma metabolomics. **A.** Volcano plot indicating the impact of COVID-19 on plasma metabolites**. B.** Sina plots with boxes indicating median and interquartile range of representative features for Kynurenine and Xanthine. q-values in each are from linear models.

**Supplementary Figure 2, related to Figures 3-5.**
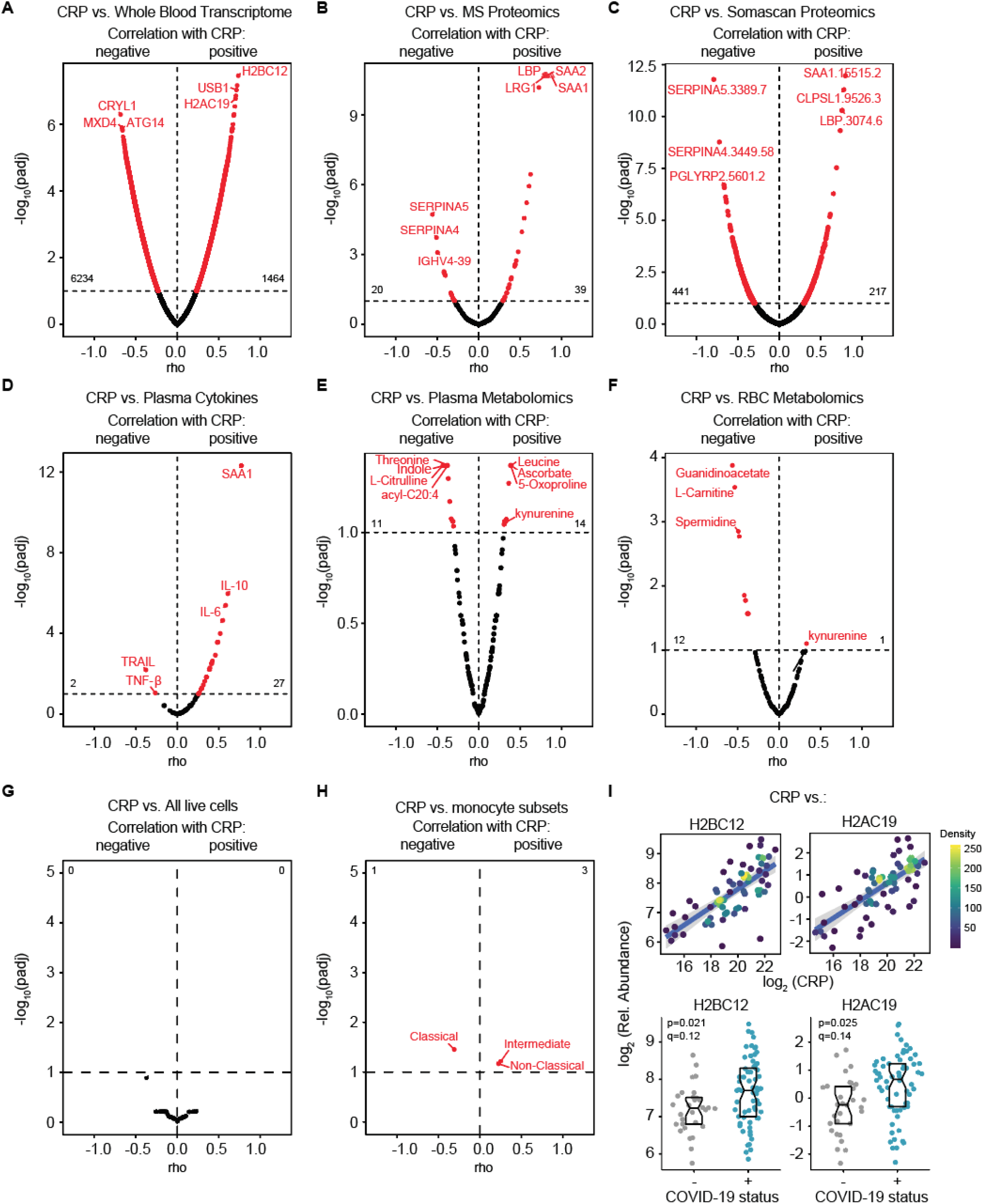
CRP correlations across the COVIDome dataset. Volcano plots for Spearman correlations between CRP levels detected by mass spectrometry (MS) and (**A**) whole blood transcriptome by RNAseq, (**B**) MS plasma proteomics, (**C**) plasma SOMAscan^®^ proteomics, (**D**) plasma cytokines, (**E**) plasma metabolomics, (**F**) red blood cell (RBC) metabolomics, (**G**), all live cell subsets detected by mass cytometry, and (**H**) monocyte subsets. The horizontal dashed lines indicated the statistical cut off of q<0.1 (FDR10). Numbers in the left and right quadrants indicate the number of features passing the statistical cut off. **I. Top**, scatter plot for correlations of CRP with indicated RNAs detected in the whole blood transcriptome. Points are colored by density; lines represent linear model fit with 95% confidence interval. **Bottom,** sina plots with boxes indicating median and interquartile range for the indicated gene. p and q values in each are from DESeq2 analysis.

**Supplementary Figure 3, related to Figure 4.**
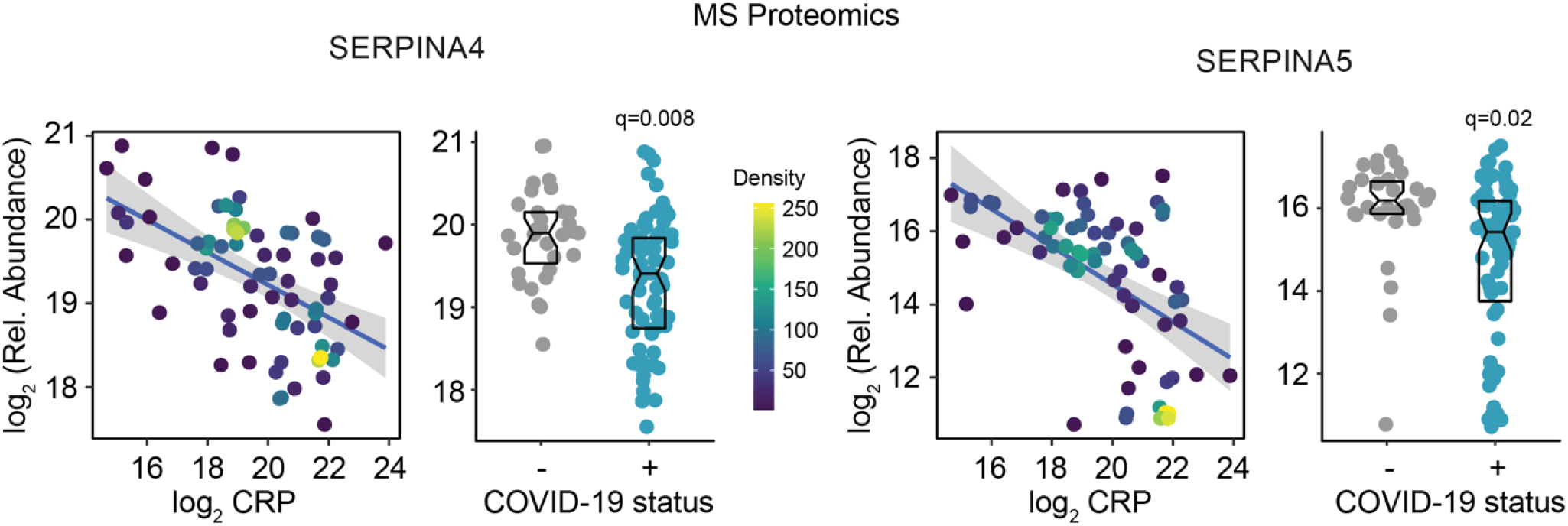
Correlation analysis of CRP with MS proteomics measurements of SERPINA4 and SERPINA5. Left, scatter plot for correlation of CRP with indicated protein. Points are colored by density; lines represent linear model fit with 95% confidence interval. q-values in each are from linear models. Sina plots with boxes indicating median and interquartile range for the indicated protein. q-values in each are from linear models.

**Supplementary Figure 4, related to Figure 5.**
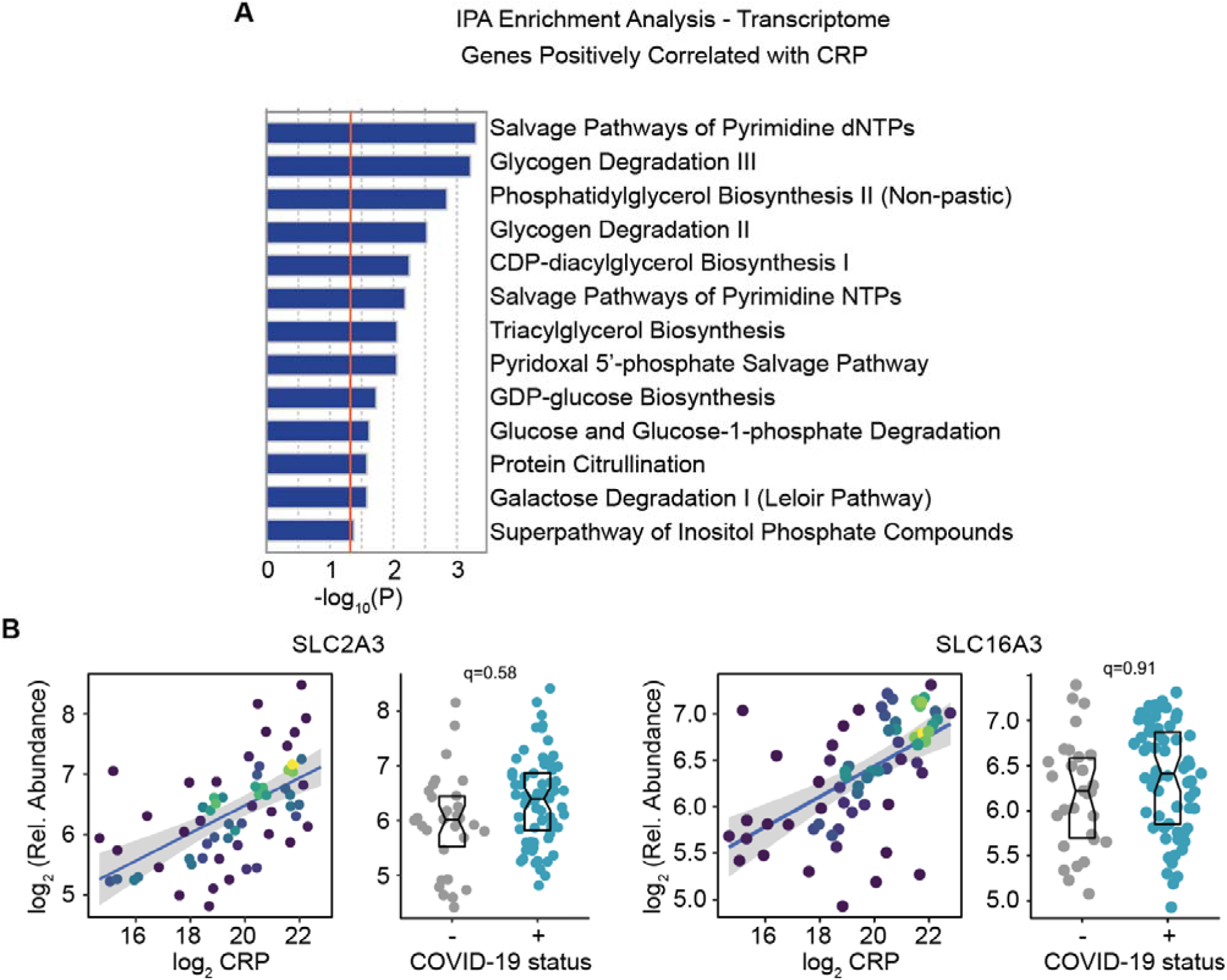
CRP positively correlates with expression of glucose and monocarboxylate transporters. **A.** Histogram displaying the results of Ingenuity Pathway Analysis (IPA) of the whole blood transcriptome for metabolic pathways enriched among mRNAs positively correlated with CRP levels. **B.** Correlation analysis of CRP with SLC2A3 and SLC16A3. **Left,** scatter plot for correlation of CRP with indicated mRNA. Points are colored by density; lines represent linear model fit with 95% confidence interval. **Right,** sina plots with boxes indicating median and interquartile range for the indicated gene. q-values in each are from DESeq2.

